# Pulmonary microbiome and transcriptome signatures reveal distinct pathobiologic states associated with mortality in two cohorts of pediatric stem cell transplant patients

**DOI:** 10.1101/2023.11.29.23299130

**Authors:** Matt S. Zinter, Christopher C. Dvorak, Madeline Y. Mayday, Gustavo Reyes, Miriam R. Simon, Emma M. Pearce, Hanna Kim, Peter J. Shaw, Courtney M. Rowan, Jeffrey J. Auletta, Paul L. Martin, Kamar Godder, Christine N. Duncan, Nahal R. Lalefar, Erin M. Kreml, Janet R. Hume, Hisham Abdel-Azim, Caitlin Hurley, Geoffrey D.E. Cuvelier, Amy K. Keating, Muna Qayed, James S. Killinger, Julie C. Fitzgerald, Rabi Hanna, Kris M. Mahadeo, Troy C. Quigg, Prakash Satwani, Paul Castillo, Shira J. Gertz, Theodore B. Moore, Benjamin Hanisch, Aly Abdel-Mageed, Rachel Phelan, Dereck B. Davis, Michelle P. Hudspeth, Greg A. Yanik, Michael A. Pulsipher, Imran Sulaiman, Leopoldo N. Segal, Birgitta A. Versluys, Caroline A. Lindemans, Jaap J. Boelens, Joseph L. DeRisi, the Pediatric Transplantation and Cell Therapy Consortium

**Affiliations:** Division of Critical Care Medicine, Department of Pediatrics, University of California, San Francisco, San Francisco, CA, USA; Division of Allergy, Immunology, and Bone Marrow Transplantation, Department of Pediatrics, University of California, San Francisco, San Francisco, CA, USA; Departments of Laboratory Medicine and Pathology, Yale School of Medicine, New Haven, CT, USA; The Children’s Hospital at Westmead, Sydney, Australia; Indiana University, Department of Pediatrics, Division of Critical Care Medicine, Indianapolis, IN, USA; Hematology/Oncology/BMT and Infectious Diseases, Nationwide Children’s Hospital, Columbus, OH, USA; CIBMTR® (Center for International Blood and Marrow Transplant Research), National Marrow Donor Program/Be The Match, Minneapolis, MN, USA; Division of Pediatric and Cellular Therapy, Duke University Medical Center, Durham, NC, USA; Cancer and Blood Disorders Center, Nicklaus Children’s Hospital, Miami, FL, USA; Harvard Medical School, Boston, Massachusetts; Division of Pediatric Oncology, Department of Pediatrics, Dana-Farber Cancer Institute and Boston Children’s Hospital, Boston, MA, USA; Division of Pediatric Hematology/Oncology, UCSF Benioff Children’s Hospital Oakland, University of California San Francisco, Oakland, CA, USA; Department of Child Health, Division of Critical Care Medicine, University of Arizona, Phoenix, AZ, USA; University of Minnesota, Department of Pediatrics, Division of Critical Care Medicine, Minneapolis, MN, USA; Department of Pediatrics, Division of Hematology/Oncology and Transplant and Cell Therapy, Keck School of Medicine, University of Southern California, Los Angeles, CA, USA; Loma Linda University School of Medicine, Cancer Center, Children Hospital and Medical Center, Loma Linda, CA, USA; Division of Critical Care, Department of Pediatric Medicine, St Jude Children’s Research Hospital, Memphis, TN, USA; CancerCare Manitoba, Manitoba Blood and Marrow Transplant Program, University of Manitoba, Winnipeg, Manitoba, Canada; Center for Cancer and Blood Disorders, Children’s Hospital Colorado and University of Colorado, Aurora, CO, USA; Aflac Cancer & Blood Disorders Center, Children’s Healthcare of Atlanta and Emory University, Atlanta, GA, USA; Division of Pediatric Critical Care, Department of Pediatrics, Weill Cornell Medicine, New York, NY, USA; Department of Anesthesiology and Critical Care, Perelman School of Medicine, Children’s Hospital of Philadelphia, University of Pennsylvania, Philadelphia, PA, USA; Department of Pediatric Hematology, Oncology and Blood and Marrow Transplantation, Pediatric Institute, Cleveland Clinic, Cleveland, OH, USA; Department of Pediatrics, Division of Hematology/Oncology, MD Anderson Cancer Center, Houston, TX, USA; Pediatric Blood and Marrow Transplantation Program, Texas Transplant Institute, Methodist Children’s Hospital, San Antonio, TX, USA; Section of Pediatric BMT and Cellular Therapy, Helen DeVos Children’s Hospital, Grand Rapids, MI, USA; Division of Pediatric Hematology, Oncology and Stem Cell Transplantation, Department of Pediatrics, Columbia University, New York, NY, USA; University of Florida, Gainesville, UF Health Shands Children’s Hospital, Gainesville, FL, USA; Department of Pediatrics, Division of Critical Care Medicine, Joseph M Sanzari Children’s Hospital at Hackensack University Medical Center, Hackensack, NJ, USA; Department of Pediatrics, St. Barnabas Medical Center, Livingston, NJ, USA; Department of Pediatric Hematology-Oncology, Mattel Children’s Hospital, University of California, Los Angeles, CA, USA; Children’s National Hospital, Washington, District of Columbia, USA; Division of Pediatric Hematology/Oncology/BMT, Department of Pediatrics, Medical College of Wisconsin, Milwaukee, WI, USA; Department of Pediatrics, Hematology/Oncology, University of Mississippi Medical Center, Jackson, MS, USA; Adult and Pediatric Blood & Marrow Transplantation, Pediatric Hematology/Oncology, Medical University of South Carolina Children’s Hospital/Hollings Cancer Center, Charleston, SC, USA; Pediatric Blood and Bone Marrow Transplantation, Michigan Medicine, University of Michigan, Ann Arbor, MI, USA; Division of Hematology, Oncology, Transplantation, and Immunology, Primary Children’s Hospital, Huntsman Cancer Institute, Spense Fox Eccles School of Medicine at the University of Utah, Salt Lake City, UT, USA; Departments of Respiratory Medicine, Royal College of Surgeons in Ireland, Dublin, Ireland; Department of Medicine, Division of Pulmonary and Critical Care Medicine, Laura and Isaac Perlmutter Cancer Center, New York University Grossman School of Medicine, New York University (NYU) Langone Health, New York, NY, USA; Department of Stem Cell Transplantation, Princess Máxima Center for Pediatric Oncology, Utrecht, Netherlands; Division of Pediatrics, University Medical Center Utrecht, Utrecht, Netherlands; Transplantation and Cellular Therapy, MSK Kids, Department of Pediatrics, Memorial Sloan Kettering Cancer Center, New York, NY, USA; Department of Biochemistry and Biophysics, University of California, San Francisco, San Francisco, CA, USA; Chan Zuckerberg Biohub, San Francisco, CA, USA

## Abstract

Lung injury is a major determinant of survival after pediatric hematopoietic cell transplantation (HCT). A deeper understanding of the relationship between pulmonary microbes, immunity, and the lung epithelium is needed to improve outcomes. In this multicenter study, we collected 278 bronchoalveolar lavage (BAL) samples from 229 patients treated at 32 children’s hospitals between 2014-2022. Using paired metatranscriptomes and human gene expression data, we identified 4 patient clusters with varying BAL composition. Among those requiring respiratory support prior to sampling, in-hospital mortality varied from 22-60% depending on the cluster (p=0.007). The most common patient subtype, Cluster 1, showed a moderate quantity and high diversity of commensal microbes with robust metabolic activity, low rates of infection, gene expression indicating alveolar macrophage predominance, and low mortality. The second most common cluster showed a very high burden of airway microbes, gene expression enriched for neutrophil signaling, frequent bacterial infections, and moderate mortality. Cluster 3 showed significant depletion of commensal microbes, a loss of biodiversity, gene expression indicative of fibroproliferative pathways, increased viral and fungal pathogens, and high mortality. Finally, Cluster 4 showed profound microbiome depletion with enrichment of Staphylococci and viruses, gene expression driven by lymphocyte activation and cellular injury, and the highest mortality. BAL clusters were modeled with a random forest classifier and reproduced in a geographically distinct validation cohort of 57 patients from The Netherlands, recapitulating similar cluster-based mortality differences (p=0.022). Degree of antibiotic exposure was strongly associated with depletion of BAL microbes and enrichment of fungi. Potential pathogens were parsed from all detected microbes by analyzing each BAL microbe relative to the overall microbiome composition, which yielded increased sensitivity for numerous previously occult pathogens. These findings support personalized interpretation of the pulmonary microenvironment in pediatric HCT, which may facilitate biology-targeted interventions to improve outcomes.

## BACKGROUND

Hematopoietic stem cell transplantation (HCT) involves high dose chemotherapy and/or radiation followed by infusion of autologous or allogeneic hematopoietic progenitor cells with the intention of correcting hematopoietic defects, rescuing chemotherapy-ablated marrow, or achieving a graft-versus-malignancy effect.^1^ HCT is often the only curative therapy for patients with life-limiting diseases such as malignancy, bone marrow failure, and inborn errors of immunity, hemoglobin, and metabolism. However, direct chemotherapy toxicity, opportunistic infection, and/or alloreactive inflammation can lead pulmonary injury in up to 40% of patients,^2–6^ which can lead to hospital mortality rates approaching 50% when mechanical ventilation is required.^7–9^

Given the severity of lung disease in this population, a deeper understanding of the pulmonary microenvironment is needed to develop next-generation diagnostic tests and treatments that will improve survival rates. The lung microenvironment is a complex interaction between pulmonary microbes, immunity, and the lung epithelium and stroma, and significant questions regarding the role of pulmonary microbes in relation to each other remain largely unanswered as they pertain to human health. We and others have shown that the lungs are not sterile, and in fact contain a variety of microbes of varying pathogenic potential that continually populate the lung due to inhalation, aspiration, and in some cases of disease, hematogenous spread.^10–12^ Lung sampling through bronchoscopic bronchoalveolar lavage (BAL) is used clinically to detect common pathogens; however, many pathogens evade detection due to preceding antimicrobial treatment, lack of serologic immunity in the post-HCT setting, or limited preselected targets on multiplex assays, all of which may lead to delayed or missed diagnoses and prolonged broad-spectrum antimicrobial exposure.^13,14^ In addition, organisms of indeterminate significance or context-dependent virulence are frequently identified, leading to questions about the structure, composition, and significance of broader microbial communities in this population.^11,15^

We previously reported that in a cohort of children preparing to undergo allogeneic HCT, both pulmonary microbial depletion and pathogen enrichment were associated with contemporaneously poor lung function, concomitant inflammation, and the eventual development of fatal post-HCT lung disease.^16,17^ To expand these findings to the post-HCT setting, we prospectively enrolled pediatric HCT patients undergoing clinically-indicated BAL as part of evaluation for pulmonary complications. BAL underwent RNA sequencing to characterize the pulmonary microbiome landscape, surveil for occult pulmonary infections, and capture lung gene expression profiles. Overall, we found that depletion of commensal microbiome constituents was associated with pathogen enrichment, acute inflammation, fibroproliferation, and poor survival. We were able to distinguish common respiratory pathogens from commensals using a community-structure analysis approach. Our results suggest a pathobiologic signature of dysbiotic lung injury that could be adapted into next-generation diagnostics and eventually leveraged in new therapeutic pipelines to improve outcomes.

## RESULTS

### Patients

From 2014-2022, pediatric HCT recipients across 32 children’s hospitals in the United States, Canada, and Australia (**Figure 1A**) who developed pulmonary complications and were preparing to undergo clinically-indicated bronchoscopic BAL were prospectively approached along with their parents/guardians for research consent to cryopreserve unused BAL (**Figure 1B**). The final cohort included n=278 BALs from n=229 patients (**Table 1**). Pulmonary symptoms developed or worsened a median 93 days after HCT (IQR 23-278) and were frequently associated with hypoxia and abnormal chest imaging, often in the setting of other comorbidities such as GVHD and sepsis (**Table 2**). BAL was performed a median 112 days after HCT (IQR 36-329), at which point lymphopenia was prevalent (median ALC 420 cells/uL, IQR 156-1,035, **eFigure 1**). Following each patient’s most recent BAL procedure, 121/229 patients required intensive care (53%), 71 required ≥7 days of mechanical ventilation (31%), and 45 patients died in the hospital (20%).

**Figure 1.**
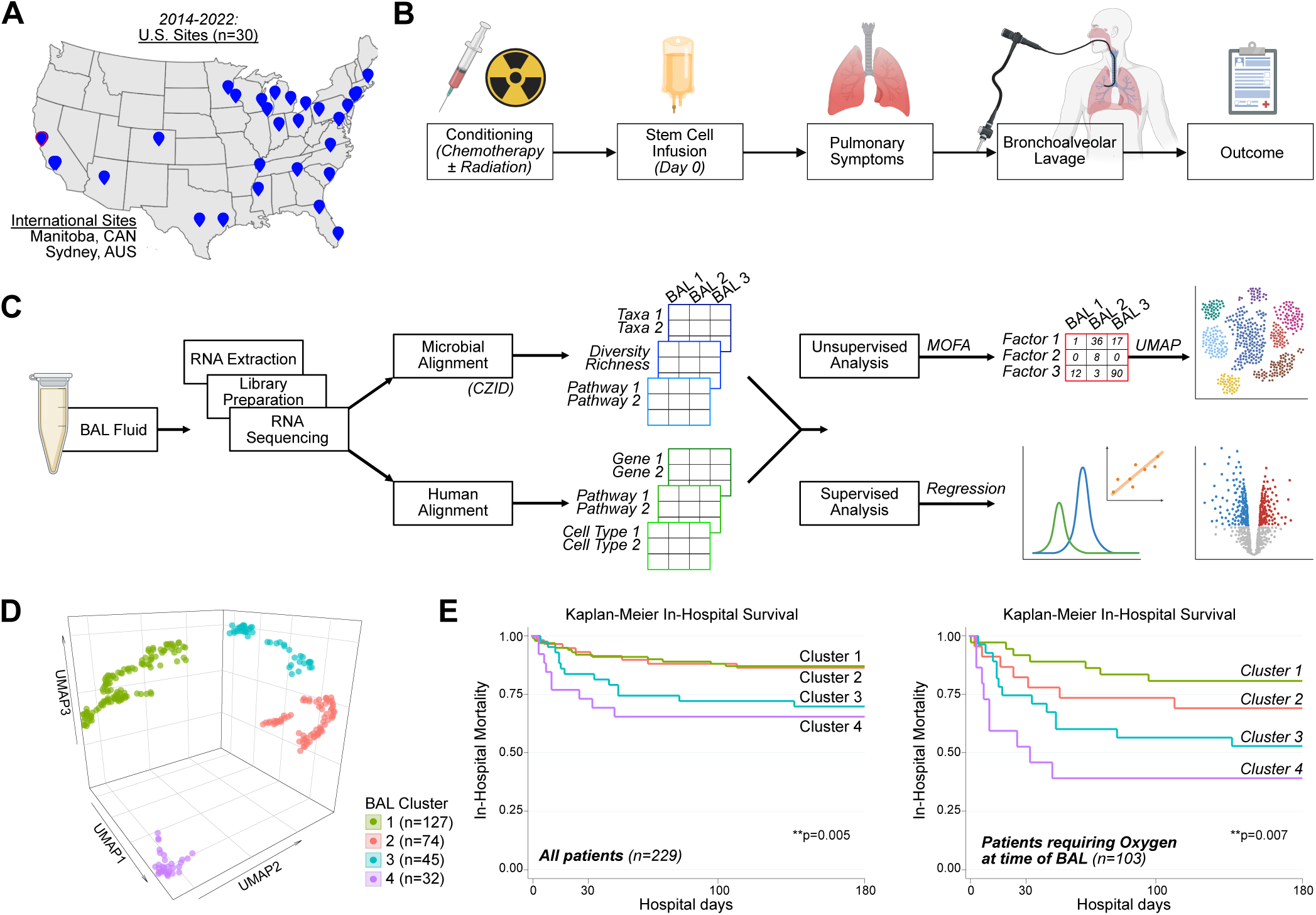
Study design and clinical outcomes. **(A)** Patients were recruited from 32 participating children’s hospitals in the United States, Canada, and Australia. **(B)** Study design concept diagram. **(C)** BAL processing and analysis workflow. **(D)** Four microbiome-transcriptome clusters were identified. **(E)** In-hospital survival for all patients (left) and the subset requiring respiratory support prior to testing (right) was plotted according to BAL cluster and differences were analyzed with the log rank test.

**Table 1.**
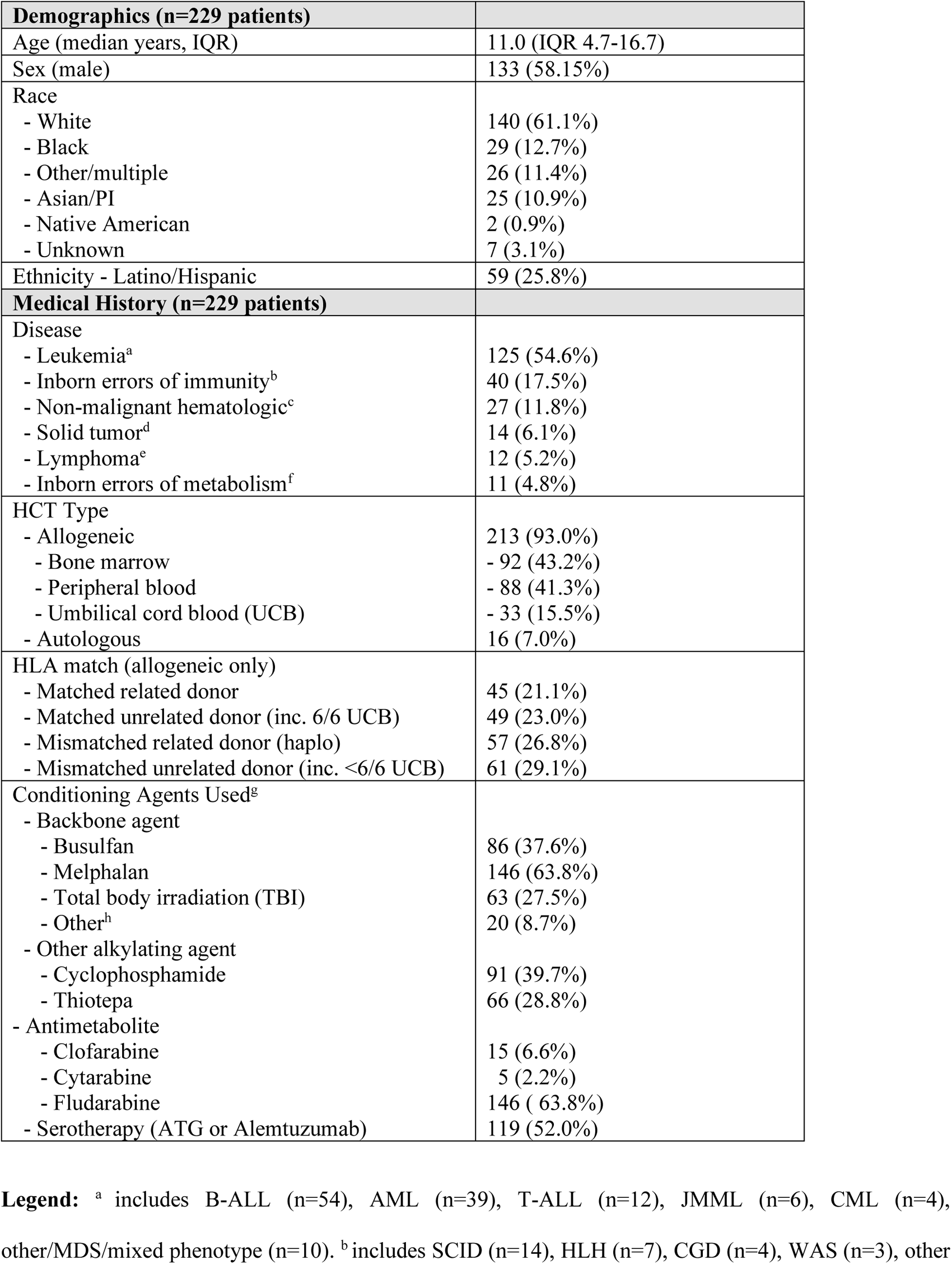
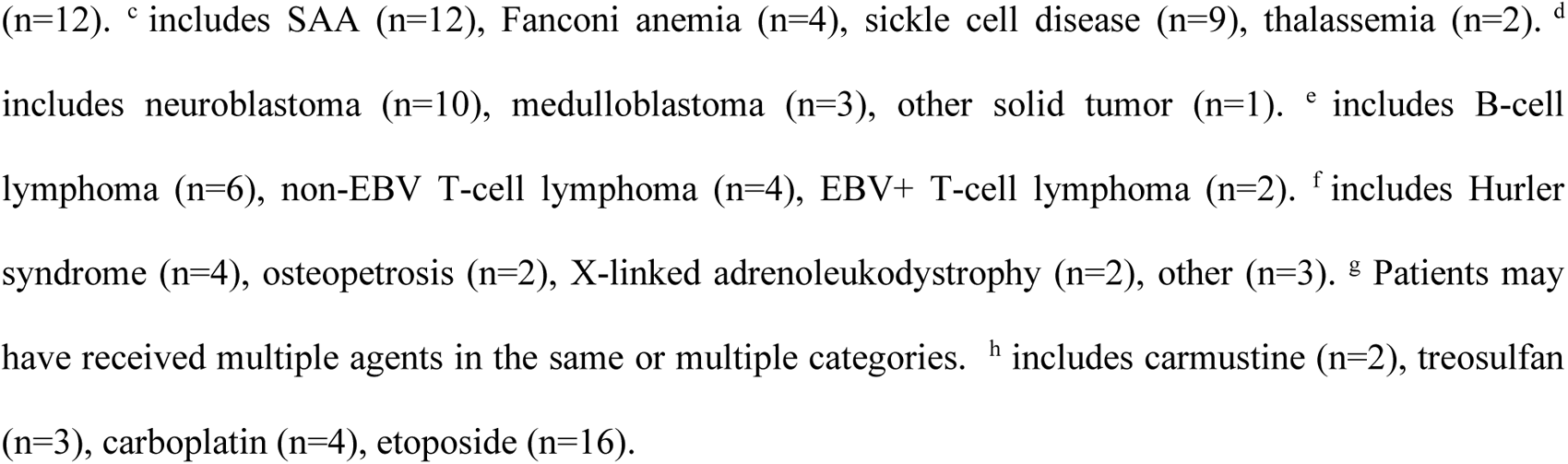
Patient Characteristics.

| **Demographics (n=229 patients)** |  |
| --- | --- |
| Age (median years, IQR) | 11.0 (IQR 4.7-16.7) |
| Sex (male) | 133 (58.15%) |
| Race |  |
| - White | 140 (61.1%) |
| - Black | 29 (12.7%) |
| - Other/multiple | 26 (11.4%) |
| - Asian/PI | 25 (10.9%) |
| - Native American | 2 (0.9%) |
| - Unknown | 7 (3.1%) |
| Ethnicity - Latino/Hispanic | 59 (25.8%) |
| **Medical History (n=229 patients)** |  |
| Disease |  |
| - Leukemia<sup>a</sup> | 125 (54.6%) |
| - Inborn errors of immunity<sup>b</sup> | 40 (17.5%) |
| - Non-malignant hematologic<sup>c</sup> | 27 (11.8%) |
| - Solid tumor<sup>d</sup> | 14 (6.1%) |
| - Lymphoma<sup>e</sup> | 12 (5.2%) |
| - Inborn errors of metabolism<sup>f</sup> | 11 (4.8%) |
| HCT Type |  |
| - Allogeneic | 213 (93.0%) |
| - Bone marrow | - 92 (43.2%) |
| - Peripheral blood | - 88 (41.3%) |
| - Umbilical cord blood (UCB) | - 33 (15.5%) |
| - Autologous | 16 (7.0%) |
| HLA match (allogeneic only) |  |
| - Matched related donor | 45 (21.1%) |
| - Matched unrelated donor (inc. 6/6 UCB) | 49 (23.0%) |
| - Mismatched related donor (haplo) | 57 (26.8%) |
| - Mismatched unrelated donor (inc. <6/6 UCB) | 61 (29.1%) |
| Conditioning Agents Used<sup>g</sup> |  |
| - Backbone agent |  |
| - Busulfan | 86 (37.6%) |
| - Melphalan | 146 (63.8%) |
| - Total body irradiation (TBI) | 63 (27.5%) |
| - Other<sup>h</sup> | 20 (8.7%) |
| - Other alkylating agent |  |
| - Cyclophosphamide | 91 (39.7%) |
| - Thiotepa | 66 (28.8%) |
| - Antimetabolite |  |
| - Clofarabine | 15 (6.6%) |
| - Cytarabine | 5 (2.2%) |
| - Fludarabine | 146 ( 63.8%) |
| - Serotherapy (ATG or Alemtuzumab) | 119 (52.0%) |

**Legend:** <sup>a</sup> includes B-ALL (n=54), AML (n=39), T-ALL (n=12), JMML (n=6), CML (n=4),
other/MDS/mixed phenotype (n=10). <sup>b</sup> includes SCID (n=14), HLH (n=7), CGD (n=4), WAS (n=3), other
(n=12). <sup>c</sup> includes SAA (n=12), Fanconi anemia (n=4), sickle cell disease (n=9), thalassemia (n=2). <sup>d</sup> includes neuroblastoma (n=10), medulloblastoma (n=3), other solid tumor (n=1). <sup>e</sup> includes B-cell lymphoma (n=6), non-EBV T-cell lymphoma (n=4), EBV+ T-cell lymphoma (n=2). <sup>f</sup> includes Hurler syndrome (n=4), osteopetrosis (n=2), X-linked adrenoleukodystrophy (n=2), other (n=3). <sup>g</sup> Patients may have received multiple agents in the same or multiple categories. <sup>h</sup> includes carmustine (n=2), treosulfan (n=3), carboplatin (n=4), etoposide (n=16).

**Table 2.**
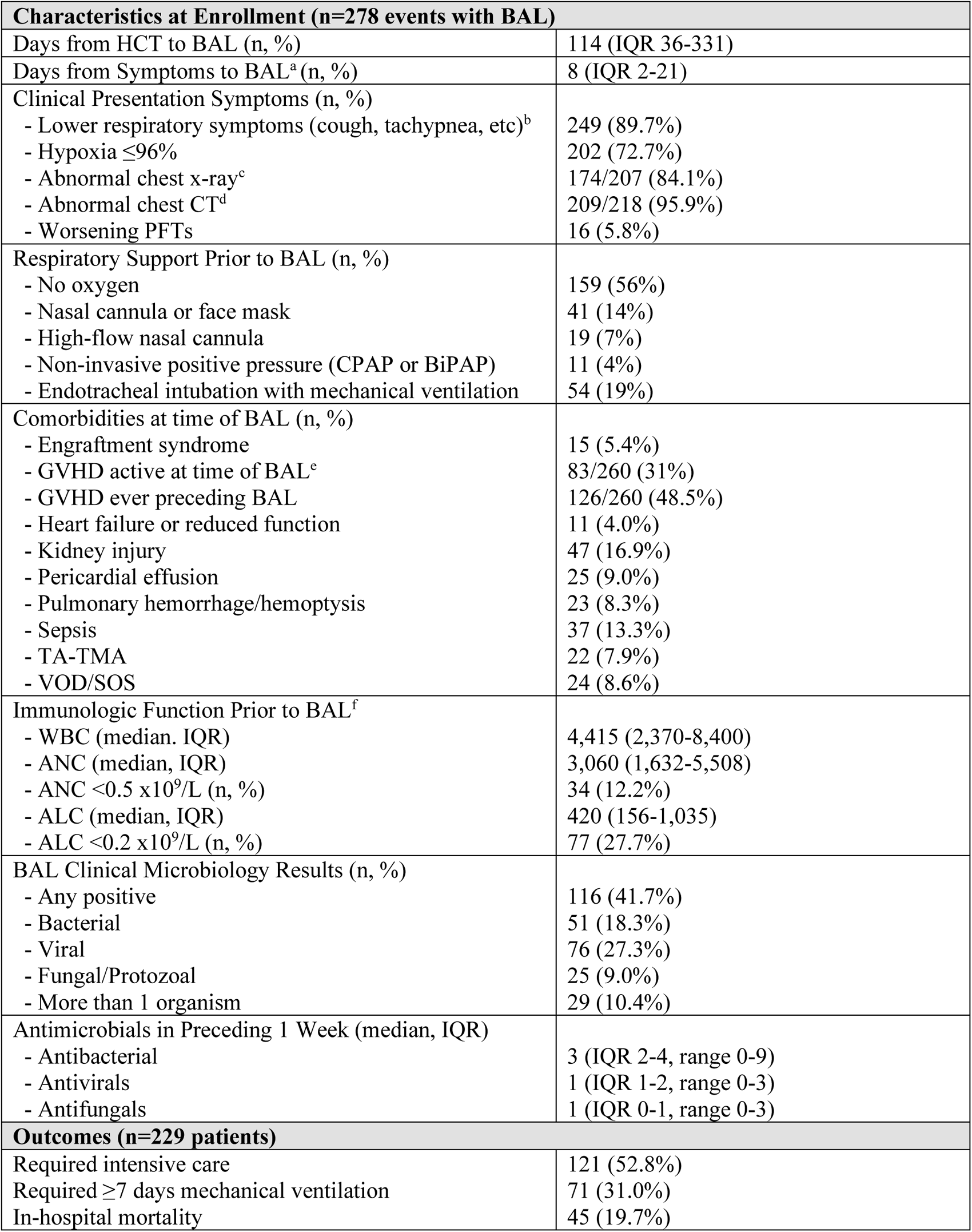
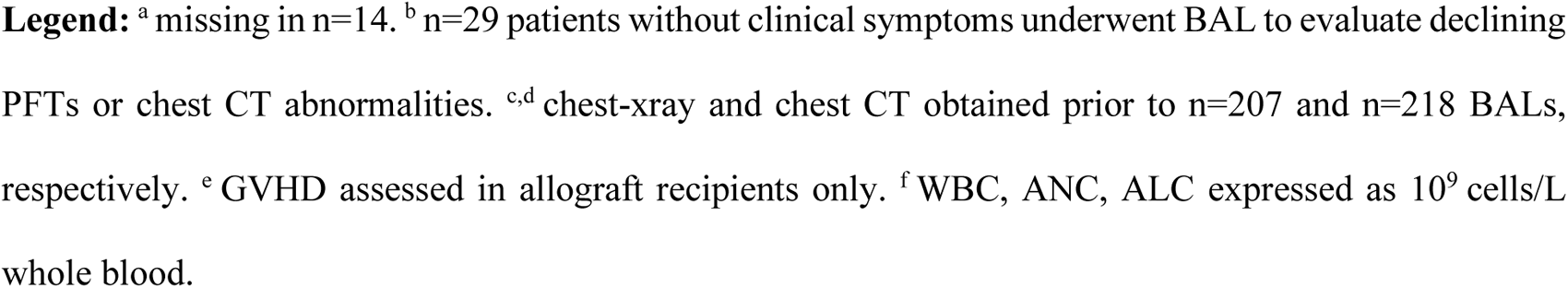
Clinical Presentation and Outcomes.

| **Characteristics at Enrollment (n=278 events with BAL)** |  |
| --- | --- |
| Days from HCT to BAL (n, %) | 114 (IQR 36-331) |
| Days from Symptoms to BAL<sup>a</sup> (n, %) | 8 (IQR 2-21) |
| Clinical Presentation Symptoms (n, %) |  |
| - Lower respiratory symptoms (cough, tachypnea, etc)<sup>b</sup> | 249 (89.7%) |
| - Hypoxia $\leq 96\%$ | 202 (72.7%) |
| - Abnormal chest x-ray<sup>c</sup> | 174/207 (84.1%) |
| - Abnormal chest CT<sup>d</sup> | 209/218 (95.9%) |
| - Worsening PFTs | 16 (5.8%) |
| Respiratory Support Prior to BAL (n, %) |  |
| - No oxygen | 159 (56%) |
| - Nasal cannula or face mask | 41 (14%) |
| - High-flow nasal cannula | 19 (7%) |
| - Non-invasive positive pressure (CPAP or BiPAP) | 11 (4%) |
| - Endotracheal intubation with mechanical ventilation | 54 (19%) |
| Comorbidities at time of BAL (n, %) |  |
| - Engraftment syndrome | 15 (5.4%) |
| - GVHD active at time of BAL<sup>e</sup> | 83/260 (31%) |
| - GVHD ever preceding BAL | 126/260 (48.5%) |
| - Heart failure or reduced function | 11 (4.0%) |
| - Kidney injury | 47 (16.9%) |
| - Pericardial effusion | 25 (9.0%) |
| - Pulmonary hemorrhage/hemoptysis | 23 (8.3%) |
| - Sepsis | 37 (13.3%) |
| - TA-TMA | 22 (7.9%) |
| - VOD/SOS | 24 (8.6%) |
| Immunologic Function Prior to BAL<sup>f</sup> |  |
| - WBC (median, IQR) | 4,415 (2,370-8,400) |
| - ANC (median, IQR) | 3,060 (1,632-5,508) |
| - ANC $< 0.5 \times 10^9/L$ (n, %) | 34 (12.2%) |
| - ALC (median, IQR) | 420 (156-1,035) |
| - ALC $< 0.2 \times 10^9/L$ (n, %) | 77 (27.7%) |
| BAL Clinical Microbiology Results (n, %) |  |
| - Any positive | 116 (41.7%) |
| - Bacterial | 51 (18.3%) |
| - Viral | 76 (27.3%) |
| - Fungal/Protozoal | 25 (9.0%) |
| - More than 1 organism | 29 (10.4%) |
| Antimicrobials in Preceding 1 Week (median, IQR) |  |
| - Antibacterial | 3 (IQR 2-4, range 0-9) |
| - Antivirals | 1 (IQR 1-2, range 0-3) |
| - Antifungals | 1 (IQR 0-1, range 0-3) |
| **Outcomes (n=229 patients)** |  |
| Required intensive care | 121 (52.8%) |
| Required $\geq 7$ days mechanical ventilation | 71 (31.0%) |
| In-hospital mortality | 45 (19.7%) |

**Legend:** <sup>a</sup> missing in n=14. <sup>b</sup> n=29 patients without clinical symptoms underwent BAL to evaluate declining PFTs or chest CT abnormalities. <sup>c,d</sup> chest-xray and chest CT obtained prior to n=207 and n=218 BALs, respectively. <sup>e</sup> GVHD assessed in allograft recipients only. <sup>f</sup> WBC, ANC, ALC expressed as 10<sup>9</sup> cells/L whole blood.

### Cluster Derivation

BAL underwent mechanical homogenization, bulk RNA extraction, and sequencing, followed by parallel alignment to microbial and human reference genomes using the open-source CZID platform (czid.org) (**Figure 1C, Methods**).^18^ Microbial alignments were transformed from reads counts to quantitative masses using a reference spike-in^19^, followed by stringent contamination subtraction^20^, and were summarized according to taxa, KEGG functional orthologs, richness, and diversity. Human alignments were characterized according to normalized gene expression, pathway analysis, cell type deconvolution, and T-and B-cell receptor alignments (**Methods**). We first used unsupervised analysis to identify underlying BAL subtypes with shared microbial-human metatranscriptomic composition. We used a two-step approach consisting of (1) multi-factor dimensionality reduction (*mofa*), followed by (2) uniform manifold approximation and projection with hierarchical clustering (*umap*) to assess BAL compositional similarity (**Methods**). Optimal fit statistics (**eFigures 2-4)** suggested that 4 clusters best fit the data structure (**Figure 1D**).

### Clinical Traits, Illness Severity and Outcomes

Clinical traits and outcomes were analyzed only after the clusters were assigned. Demographics, medical disease, transplant regimens, and graft characteristics were similar among clusters, with the exception of more females in Clusters 3 and 4 (**eTable 2**). However, patients in Clusters 3 and 4 were generally sicker, as evidenced by greater need for respiratory support prior to BAL (p=0.004), higher rates of renal injury and GVHD (p=0.001 and p=0.019), and greater use of intensive care (p=0.001) or prolonged mechanical ventilation (≥ 7 days) after BAL (p=0.001, **eTable 3**). Using each patient’s most recent BAL, patients in Clusters 3 and 4 also had significantly higher in-hospital mortality than patients in Clusters 1 or 2 (33 and 35% vs 14 and 14%, log-rank p=0.005, **Figure 1E**). Among patients requiring respiratory support prior to BAL (44%), cluster-based mortality differences were pronounced and ranged from 22-30% in Clusters 1 and 2 to 50-60% in Clusters 3 and 4 (log-rank p=0.007). Findings were similar when analyzing only patients enrolled within 100 days post-HCT (**eTable 4**) and in a multivariable Cox regression model accounting for age, biologic sex, ANC, ALC, and presence of GVHD (p=0.023, **eTable 5**). Of note, only 2 patients died within 48 hours of BAL (both in the setting of progressive septic shock).

### Microbial Taxonomy

To determine how microbiome composition drove differences between the clusters, we compared taxonomic mass, richness, and diversity. Cluster 1 was defined by moderate microbiome mass and richness, high microbial diversity, and a low burden of viruses. In contrast, Cluster 2 showed high mass of all bacterial phyla, as well as high levels of taxonomic richness and moderate microbial diversity (**Figure 2A, 2B, Data File 1).** Cluster 3 demonstrated a reduced quantity and diversity of typically oropharyngeal microbes with greater quantity of RNA viruses and the Ascomycota phylum of fungi, which contains medically-relevant pathogens such as Aspergillus, Candida, and Pneumocystis. In contrast, Cluster 4 showed significant depletion of typical microbiome constituents with minimal diversity and richness and concomitant enrichment of Staphylococcus and the Pisuviricota phylum of RNA viruses, which contains numerous respiratory RNA viruses such as Rhinovirus. BALs representative of each Cluster are shown in **eFigure 5**. We next used an orthogonal supervised analysis to compare microbiome features among survivors and non-survivors. Consistent with the description of Clusters 3 and 4, non-survivors showed broad bacterial depletion of commensal taxa, higher quantities of fungal and viral RNA (**Figure 2C, Data File 2**), and decreased BAL richness (p=0.025) and diversity (Shannon diversity p=0.006; **Figure 2D**). In contrast, survivors showed replete and bacterially diverse pulmonary microbiomes, consistent with description of Cluster 1.

**Figure 2.**
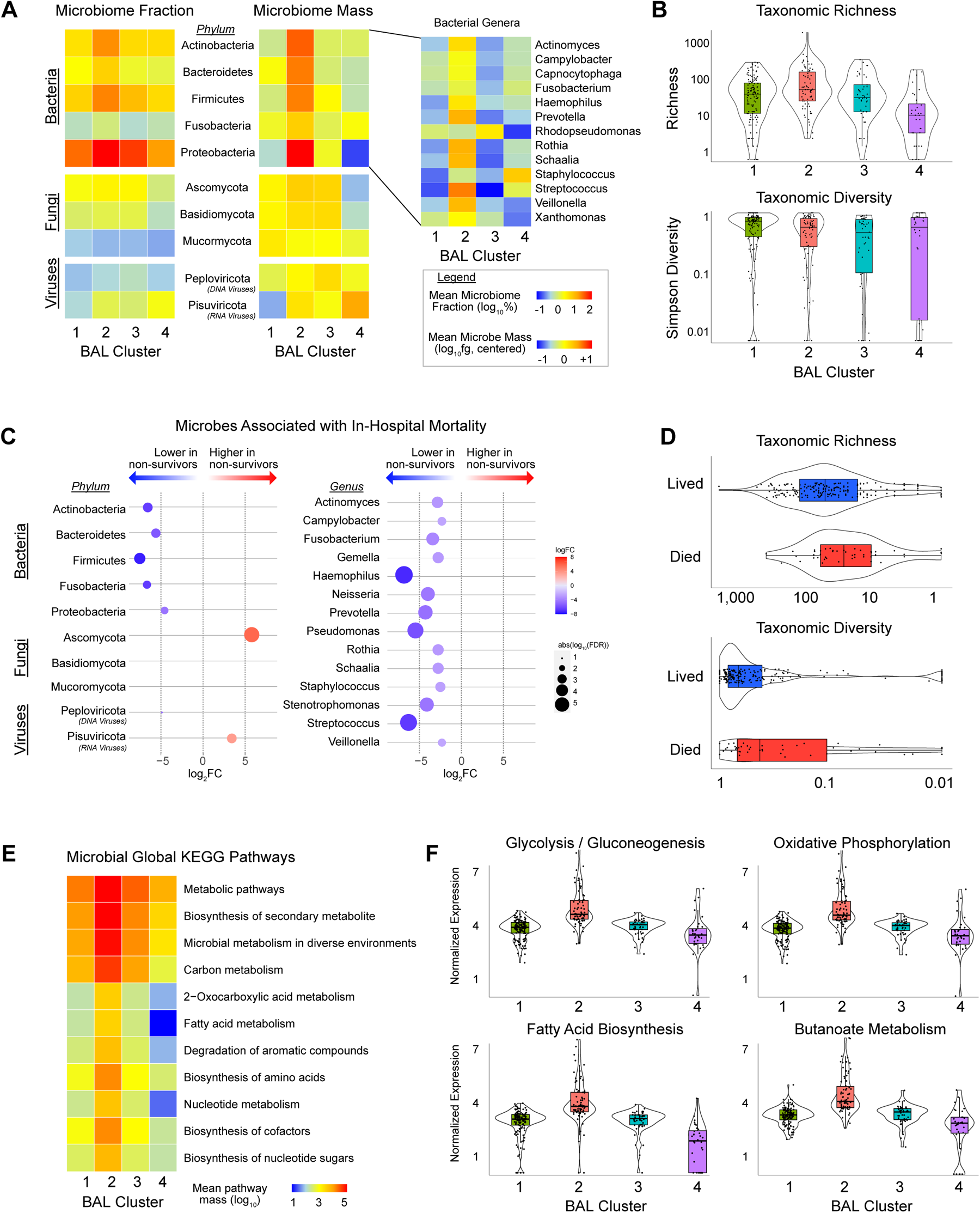
BAL microbiome. **(A)** The fraction (left) and mass (right) of major bacterial, viral, and fungal phyla are plotted, with shading representing the average for each of the 4 BAL clusters. The average mass of bacterial genera in each of the 4 BAL clusters are shown to the right. **(B)** Taxonomic richness and diversity are plotted across the 4 BAL clusters. **(C)** Microbes associated with in-hospital mortality were identified using negative binomial generalized linear models (*edgeR*) and are plotted according to logFC (position, color) and FDR (dot size). **(D)** Taxonomic richness and diversity stratified by survival status. **(E)** Microbial alignments to KEGG metabolic pathways were averaged for each BAL cluster. **(F)** Select metabolic pathways that differ across the BAL clusters are shown.

### Microbial Function

Transcriptomic markers of metabolic activity of microbial communities may complement taxonomic composition.^21^ Therefore, we next characterized the 4 clusters according to KEGG functional annotations. Cluster 1 showed moderate transcription of myriad microbial metabolic functions across the domains of carbohydrate, lipid/fatty acid, and amino acid metabolism (**Figure 2E, eFigure 6, Data File 3**). In contrast, the bacterially rich Cluster 2 showed greater transcription of these domains as well as of glycan biosynthesis pathways, including peptidoglycan, lipopolysaccharide, and other glycans that form bacterial cell walls (**eFigure 7**). Cluster 3 showed significantly lower microbial function across the spectrum of KEGG pathways, and consistent with a depleted microbiome, Cluster 4 showed minimal microbial metabolic activity. Select metabolic pathways are shown in **Figure 2F**. These results indicate that functionally, the two clusters highly associated with poor outcome showed relative loss of common critical microbial functions.

### Pathogen Identification

Although some microbiome features were shared across clusters, such as the degree of quantity of oropharyngeal taxa, many patients in this cohort had a wide range of distinct infections, thus lending unique elements to each microbiome. Therefore, we characterized the landscape of detected microbes with pathogenic potential relative to the clinical assay metadata from each patient (summarized in **eTable 6**, pathogen list in **Data File 4**, patient-level data in **Data File 5**).

#### Viruses

Clinically, most community-acquired respiratory viruses (CRVs) are detected with multiplex PCR and reported as present/absent. Clinical testing found CRVs in 18% of samples (n=49), whereas sequencing identified CRVs in 28% of samples (n=77), highest in Clusters 2, 3, and 4 (**Figure 3A**). In addition to common CRVs, several novel (< 90% nucleotide identity) or variant strains of common CRVs such as Influenza C and Rhinovirus C were detected (GenBank OQ116581, OQ116582, OQ116583).^22–24^ Clinical testing found herpesviruses (HVs) including CMV and HHV-6 in 13% of samples (n=35), whereas sequencing found HVs in 16% of samples (n=49), with greatest detection in Clusters 3 and 4. Sequencing also detected many viruses known to have respiratory transmission but not typically included on respiratory viral panels, including BK, WU, and KI Polyomaviruses, Bocavirus, Parvovirus B19, lymphocytic choriomeningitis virus (LCMV), and non-vaccine strain Rubella across 26 BALs from 23 patients. These viruses were most common in Clusters 3 and 4 and associated with 39% in-hospital mortality (n=9/23). The ubiquitous bystander torquetenovirus (TTV) and its variants were detected in 20% of samples (n=55), again higher in Clusters 2, 3, and 4 relative to Cluster 1 (**eTable 7**, p<0.001).

**Figure 3.**
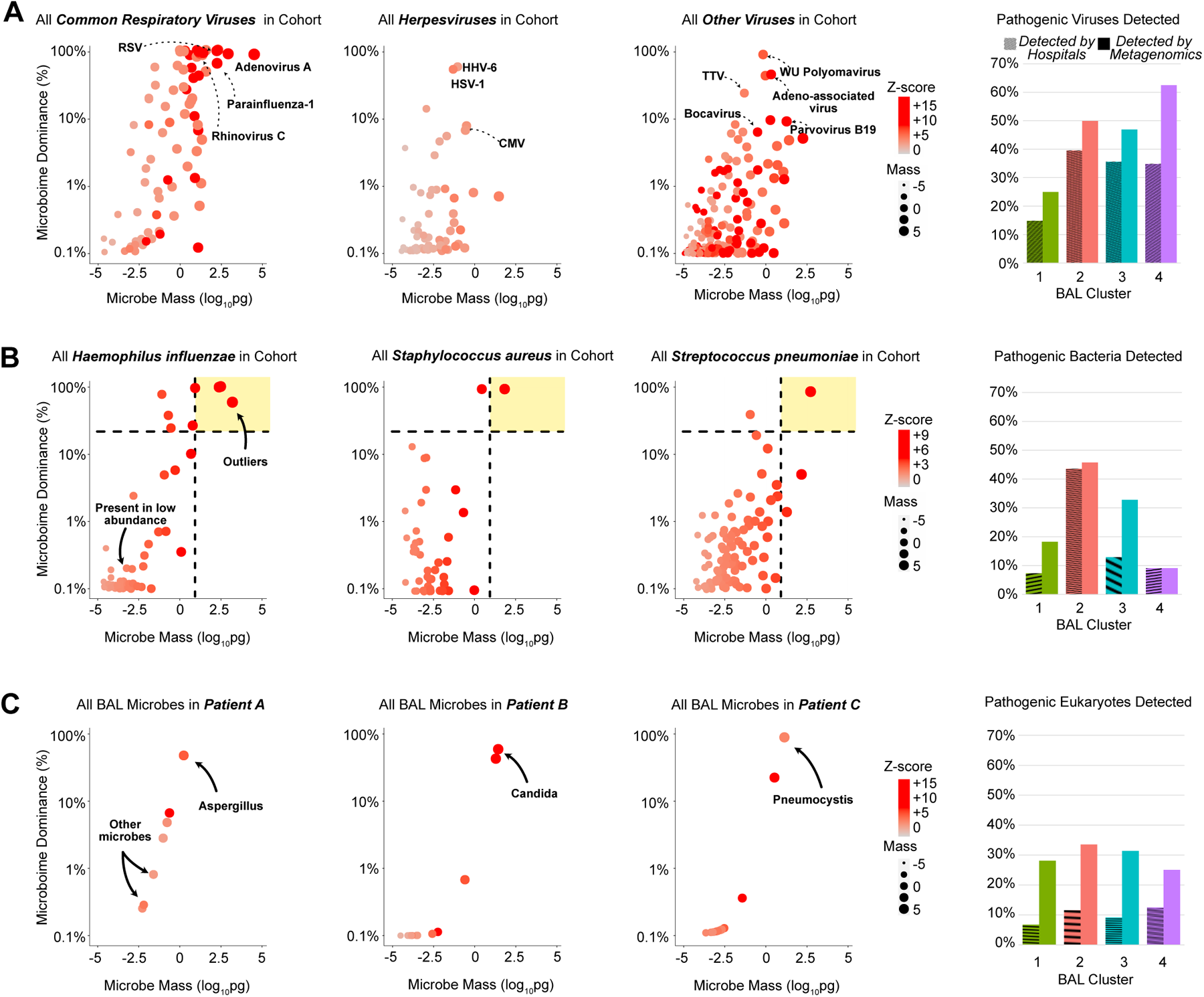
BAL pathogen detection. **(A)** Left: Dotplots of common community-transmitted respiratory viruses (left), herpesviruses (middle), and all other viruses (right) detected in the cohort, plotted according to microbial mass (x-axis) and microbiome dominance (y-axis). Right: A bar chart comparing viral detection across the 4 BAL clusters according to hospital tests and metagenomic sequencing. **(B)** Left: All *H.influenzae, S.aureus,* and *S.pneumoniae* detected in the cohort are plotted, with dotted lines indicating cutoffs of mass ≥10pg and bacterial dominance ≥20%. Taxa above these cutoffs are shown in the upper-right quadrant (shaded yellow) to indicate outliers within the cohort. Right: A bar chart comparing potentially pathogenic bacteria detected across the 4 BAL clusters according to hospital tests and metagenomic sequencing. **(C)** Left: All microbes detected in BAL of three patients are shown, with arrows pointing to fungi present in high quantities. Right: A bar chart comparing potentially pathogenic eukaryotes detected across the 4 BAL clusters according to hospital tests and metagenomic sequencing.

#### Bacteria

Clinically, most pathogenic respiratory bacteria are detected with combination of selective culture media (blood, chocolate, and McConkey agar) optimized to grow certain pathogens above non-pathogenic background, although PCR, serology, and antigen tests may be used for certain organisms. In this study, clinical testing identified pathogenic bacteria in 51 samples, which were heavily overrepresented in the microbially-rich Cluster 2 (32 of 51 bacterial infections). In contrast, metagenomic sequencing is inherently unbiased regardless of organism pathogenicity and thus can detect microbes broadly. Since contamination is ubiquitous in low-biomass samples,^25,26^ we used a strict approach to adjust for background taxa using internal spike-ins and a series of external controls (**Methods**).^20,27^ Still, many potentially pathogenic microbes were detected broadly; for example, *S.pneumoniae, M.catarrhalis, H.influenzae, S.aureus, P.aeruginosa* were detected in 34%, 21%, 21%, 16%, and 14% of samples (94, 58, 57, 44, and 39 samples), respectively. Since some microbes could be present as commensals or pathogens, depending on context and microbial burden, we then ranked bacteria according to RNA mass, dominance of the bacterial microbiome, and intra-cohort z-score in order to parse microbes most likely to be present in states of dysbiosis and thus potential infection (**Figure 3B**). Using a conservative threshold of RNA mass ≥10pg, bacterial dominance ≥20%, and Z-score ≥+2, we found potentially pathogenic bacteria in 76 samples, again with nearly half of these in Cluster 2. In addition to new cases of common pathogens (e.g.: *P.aeruginosa*), numerous previously occult pathogens were identified above these thresholds, including *B.cereus, C.freundii, C.pneumoniae, K.aerogenes, S.enterica,* and *U.parvum*.

#### Eukaryotes

As with bacteria, many potentially pathogenic fungi were detected broadly in this cohort; for example, Candida, Aspergillus, Fusarium, and Rhizopus were detected in 18%, 16%, 9%, and 5% of samples (50, 44, 25, and 13), respectively. By clinical assays, potentially pathogenic fungi were detected in 9% of samples (n=25). Using sequencing with a threshold of mass ≥10pg and Z-score ≥+2, potentially pathogenic fungi were detected in 30% of samples (83), with high detection across clusters 2, 3, and 4 (**Figure 3C**). Several relevant fungi were detected exclusively by metagenomic sequencing, including Cryptococcus and Pneumocystis. No BAL parasites were detected through clinical assays, whereas metagenomic sequencing detected Toxoplasma in 4 patients and Acanthamoeba in 3 patients, with predominance in Clusters 3 and 4 (**Data File 5**) and >50% mortality rate (n=4/7).

Overall, clinical testing identified 173 pathogens in 116/278 samples (41.7%), while metagenomic sequencing using the above conservative thresholds identified 360 pathogens in n=196/278 samples (70.5%, McNemar’s p<0.001, **eTable 8**). Combined, clinical testing and metagenomic sequencing together identified 429 pathogens in n=209/278 samples (75.2%, **eTable 6**). Whereas clinical testing identified pathogens in 22/45 non-survivors (49%), sequencing identified credible pathogens in 36/45 non-survivors (80%, p=0.002). In-hospital mortality was highest for those with a pathogen detected by both clinical testing and metagenomics and lower if a pathogens was detected by metagenomics alone or was not detected at all (27% vs 19% vs 13%, **eTable 9, eFigure 8**).

### Impact of Antimicrobial Exposure

Although the effects of antimicrobial exposure have been demonstrated on the intestinal, nasal, and oropharyngeal microbiomes, the effects of antibiotics on the bronchoalveolar microbiome are less clear, with some reporting a major effect^28–35^ and some reporting minimal effect.^36,37^ To investigate this, we quantified patient-level antibacterial exposure in the week preceding BAL by weighting the cumulative antibiotic exposure days (**Figure 4A**) with an agent-specific broadness score^38^ to yield an antibiotic exposure score (AES, **Figure 4B, Methods**). AES varied across clusters (p=0.005) and was lowest for the microbially-rich Cluster 2 and highest for the microbially depleted Clusters 3 and 4. Greater AES was associated with reduced BAL microbial richness (Spearman rho -0.14, p=0.018); depletion of all the major bacterial phyla including numerous oropharyngeal-resident taxa; and enrichment of the fungal phylum Ascomycota (FDR<0.05, **Figure 4C, Data File 6**). In addition, AES was significantly greater among non-survivors (median 352, IQR 210-507 vs. 175, IQR 75-336, Wilcoxon rank-sum p<0.001), with sequentially higher mortality with increasing AES quartile (**eFigure 9**). Using causal mediation analysis based on linear structural equation modeling (**Methods**), the association between greater AES and mortality was statistically mediated by an antibiotic-induced reduction in key commensal pulmonary bacteria including Actinomyces, Fusobacterium, Gemella, Haemophilus, Neisseria, Rothia, Schaalia, and Streptococcus (p<0.001), suggesting that the link between antibiotic exposure and mortality can at least partially be explained by effects of antibiotics on the pulmonary microbiome (**eFigure 10, Data File 7**). Many groups have found that anti-anaerobe exposure is associated with a depleted intestinal microbiome and progression of upper respiratory viral infections to the lower respiratory tract.^39–41^ Similar to above, anti-anaerobic exposure was higher in non-survivors (p=0.011) and was associated with BAL depletion of numerous anaerobes including Prevotella, Gemella, and Fusobacterium (**Data File 8**). Anti-fungal exposure appeared higher in the microbially-depleted Cluster 4, driven largely by higher exposure to echinocandins (p=0.019), and anti-viral exposure appeared higher in Clusters 3 and 4, driven largely by higher exposure to cidofovir (p=0.045).

**Figure 4.**
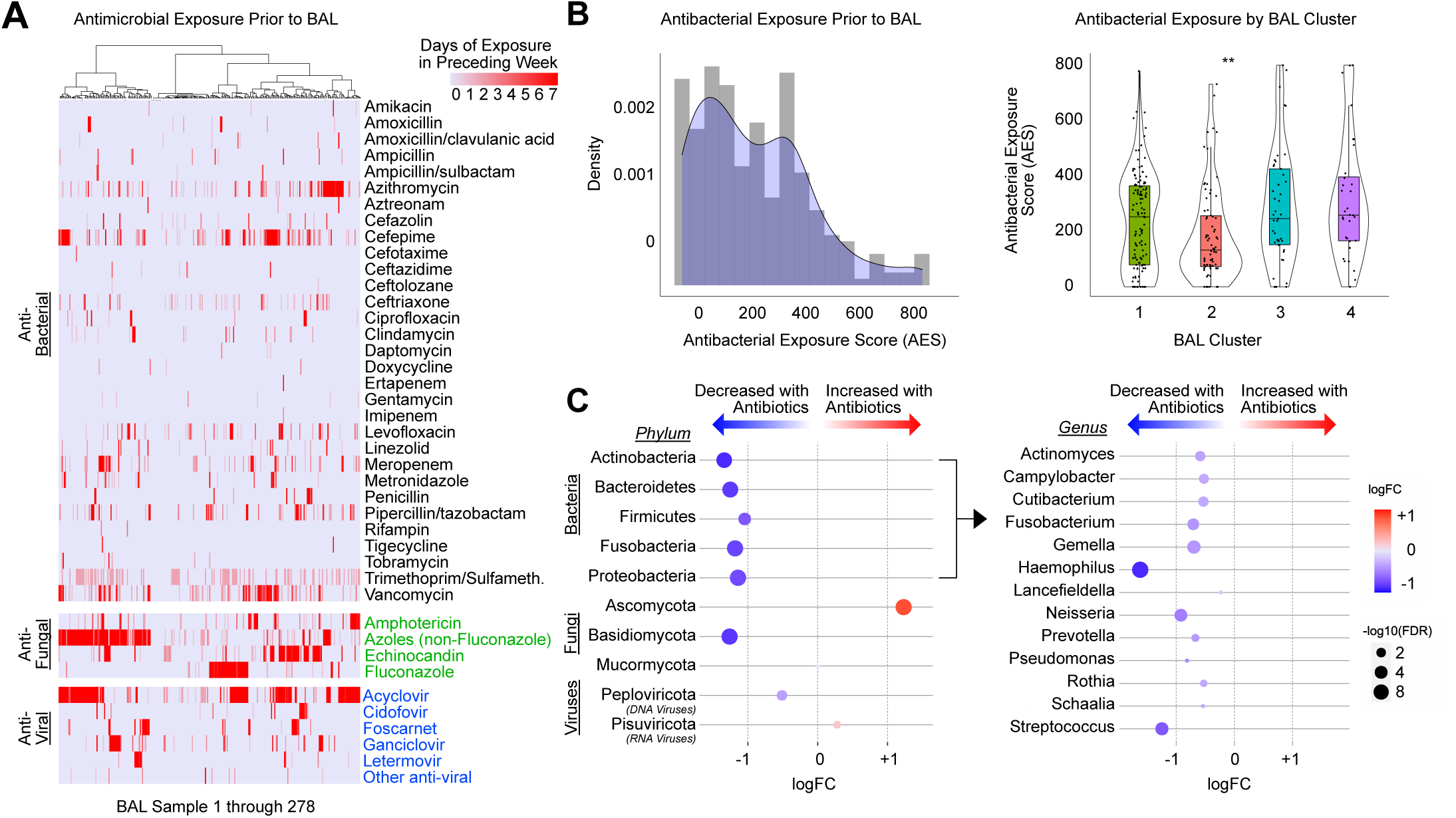
Antibiotic exposure and impact on BAL microbiome. **(A)** Days of antimicrobials are listed for antibacterials (black), antifungals (green), and antivirals (blue). Patients are listed in columns and shading indicates number of days of exposure to each antibiotic in the week preceding BAL. **(B)** Antibiotic exposure score (AES) was calculated prior to each BAL as the sum of antibiotic exposure days times a broadness weighting factor, summed for all therapies received in the week preceding BAL. AES varied across the clusters and was highest for patients in Cluster 4. **(C)** Negative binomial generalized linear models were used to test for BAL microbes associated with AES. Microbes are listed in rows, with phyla shown on the left and bacterial genera shown on the right.

### Impact of Clinical Immune Status

The pulmonary microbiome exists in a state of reciprocal interaction with the lung epithelium, stroma, and immune system. To contextualize microbiome states according to systemic immunity, we analyzed each patient’s most recent blood absolute neutrophil count (ANC) and absolute lymphocyte count (ALC) measured prior to BAL. ANC was highest in the bacterially-rich Cluster 2 (p=0.029, **eTable 3**) but was not associated with mortality overall (p=0.810). ALC did not vary across clusters (p=0.997) but was significantly lower in non-survivors (median 273, IQR 125-650 vs. 422, IQR 179-1120, p=0.028).

### Pulmonary Gene Expression

We then compared BAL human gene expression across the 4 clusters. A 4-way ANOVA-like analysis yielded 18,158 genes differentially expressed across the 4 clusters (**Figure 5A, Data File 9**). Select genes most differentially expressed in each cluster are displayed in **Figure 5B**. To assess the biological pathways represented by these genes, we compared GSVA enrichment scores for Reactome gene sets (**Data File 10**); select pathways most differentially expressed in each cluster are displayed in **Figure 5C**. Overall, Cluster 1 showed high expression of pathways related to antigen-presenting cell activation; Cluster 2 showed high expression of genes and pathways related to neutrophil and innate immune activation, bacterial processing, and airway inflammation; Cluster 3 showed high expression of pathways related to collagen deposition and fibroproliferation; and Cluster 4 showed high expression of anti-viral and cellular injury genes. To replicate these findings using a different methodology unrelated to the above clusters, we performed a supervised analysis comparing gene expression among survivors and non-survivors and identified 1,253 differentially expressed genes (**Data File 11**). Consistent with the description of Clusters 3 and 4, BALs from non-survivors showed broad down-regulation of innate immune and antigen-presenting signals and a significant upregulation in collagen deposition, matrix metalloproteinases, alveolar epithelial hyperplasia, and fibroproliferative genes (e.g.: COL1A1, COL3A1, CXCL5, IL13, MMP7, SFTPA1, SFTPC, TIMP3).

**Figure 5.**
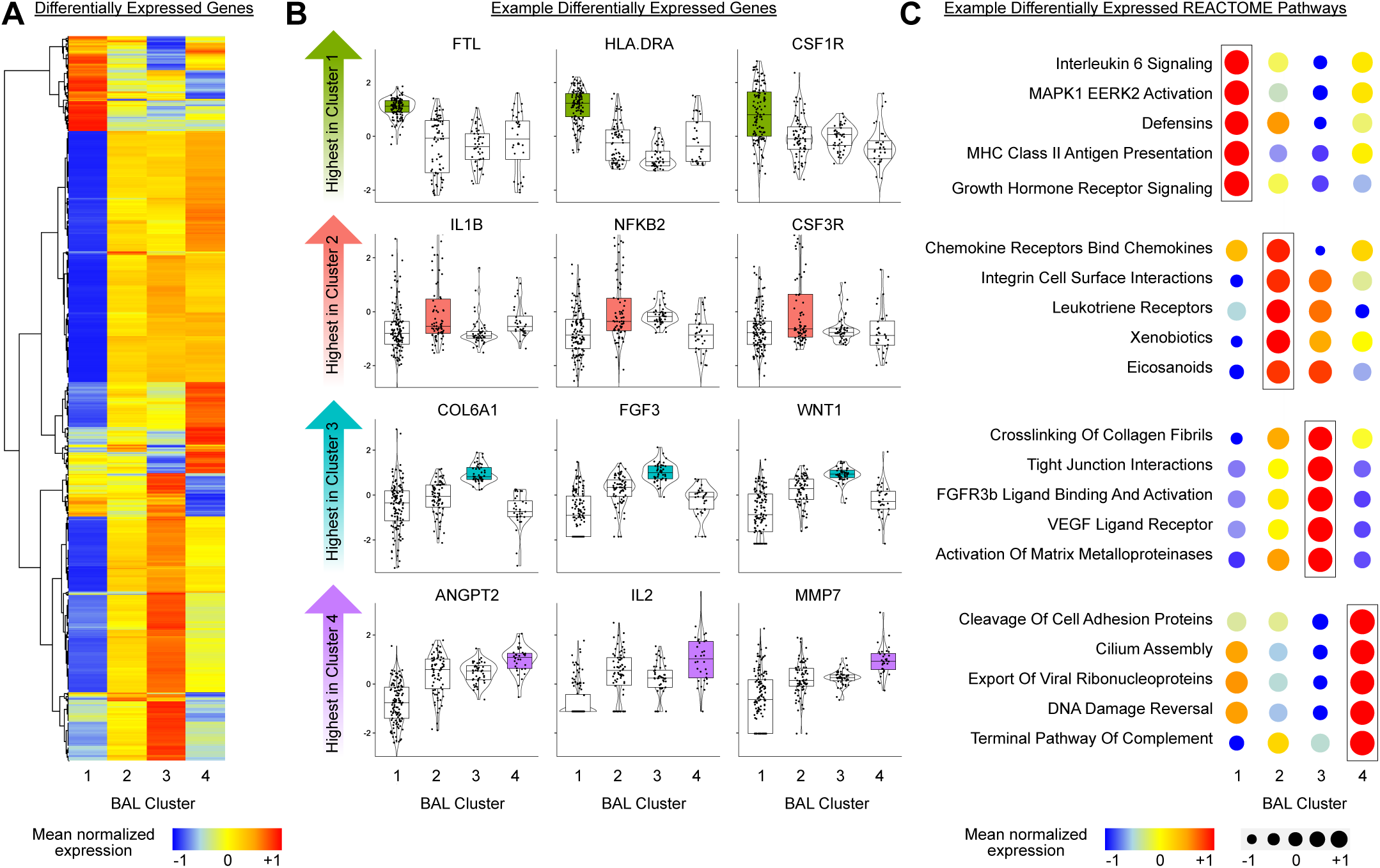
BAL gene expression. **(A)** Differentially expressed genes were identified by 4-way ANOVA like analysis with negative binomial generalized linear models. Mean normalized expression levels for significant genes are displayed for the 4 BAL clusters. **(B)** Individual differentially expressed genes were identified across the 4 clusters (*edgeR*) and variance-stabilized transformed gene counts for select genes highest in each of the 4 clusters are plotted. **(C)** Gene set enrichment scores to Reactome pathways were calculated and example gene sets most enriched in each of the 4 clusters are shown.

### BAL Cell Type Imputation

BAL contains an admixture of cell types in contact with the lumen of the lower respiratory tract, and thus varying cell proportions may account for differential gene expression detected by bulk sequencing. A multi-center study with small volume of BAL samples precluded single cell sequencing. Hence, we used *in silico* cell type deconvolution with CIBERSORTx and the *Travaglini* lung cell atlas to impute cell fractions in each sample.^42–44^ Consistent with findings described above, Cluster 1 showed high representation of antigen presenting cells including monocytes and macrophages, Cluster 2 showed a greater fraction of neutrophils, Cluster 3 showed a paucity of innate immune cells and a higher fraction of CD4+ T-cells, and Cluster 4 showed a high fraction of CD8+ T-cells (**eFigure 11**). Given the findings of varying cellular fraction within the BAL clusters, we then imputed cell-type specific gene expression using CIBERSORTx (**Methods**). Monocyte-specific expression of the GOBP “Myeloid Leukocyte Activation” gene set varied across clusters, with higher activation of activation markers such as CSF1, IFNGR1, LDLR, TLR1, and TNF seen in Clusters 2, 3 and 4; notably, although Cluster 1 had a high monocyte/macrophage cell fraction, lineage-specific inflammatory gene activation was relatively low in this cluster (**eFigure 12**). Similarly, lymphocyte-specific expression of the GOBP “Lymphocyte Activation” gene set varied across clusters, with the highest levels of markers such as AKT1, BTK, CD4, DOCK8, JAK2, and IL7R seen in Clusters 3 and 4 (**eFigure 13**). Given the varying cell proportions and imputed activation levels of lymphocytes across the clusters, we next aimed to determine whether there might be differences in lymphocyte repertoires across the clusters. Using ImRep, we identified that the majority of CDR3 alignments were for TCRα, with many fewer alignments to β, γ, and δ as well as to BCR H, K, or L. Whereas the virally-enriched Cluster 4 showed the highest number of TRA clonotypes and diversity, Cluster 1 showed the lowest (**eFigure 14**). Notably, BAL TCRαβ clonotype numbers and diversity were not correlated with blood lymphocyte count (p=0.646), although BAL TCRγδ and BCR subtypes were higher in patients with higher blood ALC (p=0.041 and p=0.006, respectively).

### Cluster transitions

We next assessed whether original cluster assignments were stable over time. Thirty-four patients had ≥2 BALs separated by a median of 79 days (IQR 21-243). Most patients who started in the low-risk Cluster 1 moved out of Cluster 1 (17/26) to a higher-risk cluster, and patients who started outside of Cluster 1 rarely moved into Cluster 1 (8/49), driving an overall change in the cluster burden over time (p<0.001, **eFigure 15, eTables 10-11**). This suggests that, for the subtype of patients with recurrent or non-resolving lung disease, progression to an adverse BAL phenotype is common over time.

### Classification Model and External Cluster Validation

Finally, as cluster assignments cannot be directly applied to external cohorts, we used taxonomic and gene expression data to grow a random forest of 10,000 classification trees with a maximum depth of 10 nodes to be used as a cluster classifier. Out-of-bag AUC was 0.923 indicating good cluster discrimination (**eTable 12**). Lung gene expression variables were significantly more important to cluster classification than were taxonomic variables, with the 500 most important genes showing significant enrichment for immune processes (**Data Files 12-13**). The random forest classifier was then applied to taxonomic and gene expression data from an independent cohort of n=57 BALs obtained from pediatric HCT recipients at the University Medical Center in Utrecht, the Netherlands, between 2005-2016 (clinical traits described in **eTable 13**). Although this cohort differed in geography, underlying diseases, allograft characteristics, and treatment protocols, 1-year non-relapse mortality was lowest among patients with BALs assigned to the low-risk Cluster 1 (9%, 2/21), was higher for patients assigned to the bacterially-rich Cluster 2 (36%, 4/11), and was highest for patients in the high-risk Clusters 3 or 4 (52%, 13/25, p=0.009, **eFigure 16, eTable 14**), thus confirming the external validity and clinical significance of the BAL cluster profiles.

## DISCUSSION

Lung injury in pediatric hematopoietic cell transplant patients is frequently fatal, yet a lack of investigable biospecimens has hindered progress in elucidating the pathobiology of disease. In this prospective multicenter study, we used BAL from children at 32 hospitals to identify microbe-lung transcriptomic signatures shared across patients. Although each BAL archetype was associated with undue morbidity, microbial dysbiosis, undetected infection, and subtypes of inflammation and fibroproliferation were identified as the primary hallmarks of fatal disease. Our findings come from a broad, international cohort of children with poor immunity and high antimicrobial exposure and were replicated in an unrelated validation cohort, thus lending credence to the work. These findings extend our previous work in pediatric HCT candidates and suggest the possibility for precision pulmonary phenotyping as a key first step for future interventional trials.

A major finding of our work is the identification of heterogeneous disease biology within a cohort of medically complex patients where disease classification has been historically difficult.^2^ BAL Cluster 1 was most common, had moderate microbial burden, low rates of infection, predominantly alveolar macrophage-related signaling, and the lowest mortality rates. In contrast, Cluster 2 showed high rates of microbial burden and bacterial infections, higher blood neutrophil counts and BAL neutrophil-related gene expression, and moderate mortality. Cluster 3 showed general microbiome depletion with enrichment of viruses and fungi and fibroproliferative gene expression. Cluster 4 showed significant microbiome depletion with relative sparing of Staphylococci and enrichment of viruses, commensurate with lymphocytic inflammation, cellular injury, and the highest mortality rate. In the field of pulmonology, subclasses of asthma, acute respiratory distress syndrome, and chronic obstructive pulmonary disease have recently been associated with distinct clinical trajectories such that subclass-specific clinical trials are now emerging.^45–47^ The identification of heterogeneous clusters may be the first step in improving bedside phenotyping and ultimately enrolling pediatric HCT patients in biology-targeted interventional trials.

A second major finding of our work is the illumination of the delicate balance between the pulmonary microbiome and mortality. The pulmonary microbiome is populated early in life by aerosolization of oropharyngeal microbes during tidal ventilation, gastric aspiration, and disease-related hematogenous spread.^10,12,48,49^ The near continuous exposure of the lungs to microbes introduces the opportunity for infection but also supports immune and epithelial education in the form of tolerance and memory.^50,51^ The ideal properties of the peri-HCT pulmonary microbiome likely require delicate balance between over-population and eradication.^10,12^ Favoring the former, studies in cystic fibrosis and COPD have shown that an increase in pulmonary microbial mass is associated with neutrophilic inflammation and disease exacerbations^52–55^, a paradigm similar to patients in our bacterial- and neutrophil-enriched Cluster 2. Favoring the latter, recent studies show that HCT patients with depleted or dysbiotic intestinal microbiomes develop higher mortality rates due to excess colitis, graft versus host disease, and even pulmonary disease, which is similar to patients in our Clusters 3 and 4.^56–58^ Our data show that a biodiversity and richness exist reciprocally with pathogenic taxa such as *S.aureus, P.aeruginosa*, fungi, and viruses, suggesting that commensal constituents may limit the ability for pathogens to expand,^59,60^ perhaps through local immunomodulation or by direct nutrient competition.^54,61–65^ We show that the transcriptional activity of BAL microbes is quite broad in patients with better clinical outcomes, raising the possibility that microbial metabolites might benefit airway health, as has been recently shown for the anti-apoptotic microbial metabolite indole-3-acetic acid (IAA).^21,66,67^

Given the findings of commensal microbial depletion in non-survivors, we explored potential exposures leading to this state. Antimicrobial exposure, the most likely culprit, has been strongly associated with intestinal microbiome depletion and to a lesser extent pulmonary microbiome alterations mostly in the cystic fibrosis and COPD populations.^28–35^ Our unsupervised analysis showed the highest antibiotic exposure in the most bacterially-depleted BAL cluster 4, which complements the supervised analysis finding of a negative relationship between AES and the quantity of numerous commensal bacteria. Causal mediation modeling showed that association between AES and death was largely mediated by antibiotic-induced contraction in BAL bacteria. Interestingly, we found that the quantity of the fungal phyla Ascomycota increased with greater AES, supporting existing evidence that depletion of commensal microbes may open a niche for opportunistic fungal growth.^68–71^ Increased AES was associated with greater BAL quantity of respiratory RNA viruses, consistent with previous associations between antibiotic exposure and viral expansion.^40,72^ Although other factors such as the conditioning regimen may influence microbiome composition,^73^ these data argue for the need for judicious use of antibiotics, which might best be achieved with rapid turnaround of clinical metagenomics assays in the future.^74^ Certainly for critically ill patients with unclear diagnoses, it will be difficult to feel confident in stopping antibiotics. Therefore, microbiome-restorative therapies in patients necessarily antibiotic-exposed may be a crucial tool.^75^

Over the past thirty years, numerous studies have confirmed that metagenomic sequencing for a wide range of indications, such as meningitis and encephalitis, can increase diagnostic yield for pathogens.^76–78^ However, application of metagenomics to respiratory fluid has been hindered by difficulty discriminating when a normal microbiome constituent such as *S.pneumoniae* expands to function as a pathogen. To address this, we transformed our sequencing data from fractional to absolute using reference spike-ins and then compared each microbe’s detected level to that of other microbes in the sample (dominance) as well as to other samples in the cohort (z-score). By parsing microbes in the context of the broader microbiome, we provide a logical and intuitive approach to pathogen detection in non-sterile body sites. This approach nearly doubled the number of patients with detected infections, while also providing a safeguard against overcalling hits. Importantly, we identified novel viral strains, common and rare bacteria, and numerous fungi and parasites as previously undetected causes of lung injury. Pathogen detection was highest in the most dysbiotic clusters with the greatest commensal depletion and lowest richness and diversity, lending credence to the above findings that airway commensals may safeguard the lungs against opportunistic infections. Our data support the premise of a clinical trial using metagenomics to augment the clinical utility of hospital diagnostics specifically in the setting of HCT. While many newly detected microbes have existing effective treatments, many lack therapies at this time. Given our findings implicating antimicrobial exposure with dysbiosis and poor clinical outcomes, antibiotic de-escalation or avoidance of dysbiosis may be useful outcome metrics for such a trial.

The relationship between the pulmonary microbiome, lung epithelium, and the transplanted immune system is characterized by a continuous and mutually influential interaction. In murine models of allogeneic HCT, immune responses to pathogens can be both impaired as well as exaggerated, leading to delayed phagocytosis, excessive myeloid cell recruitment and unremitting inflammation due to a lack of functional NK-and T-cells.^79–82^ Our data support this paradigm in human patients and reveal a complex and heterogeneous immune response. Cluster 1, with a replete and diverse pulmonary microbiome, showed the lowest mortality rates, low levels of granulocyte activation, and low levels of lymphocyte diversity and lymphocyte-specific activation markers. In contrast, Cluster 2 showed neutrophil enrichment, and Clusters 3 and 4 showed a diverse lymphocyte population with markers of activation. Clinically, these distinctions may be important, as patients might benefit from different approaches to immunomodulation. Notably, Cluster 3 showed numerous markers of fibroproliferation and cellular senescence, suggesting transition to a fibrotic phenotype that may merit treatment in upcoming clinical trials using novel anti-fibrotic agents.^83^

This study has several limitations. First, the cohort’s clinical heterogeneity requires interpreting findings broadly. Second, clinical protocols were not standardized and thus varying post-HCT care across centers could have influenced outcomes. Third, BAL collection was not standardized across centers and bronchoscope controls were not obtained. However, our approach to adjusting for contamination used ample internal and external controls. Fourth, detailed concurrent immunosuppressive regimens were not collected. Fifth, despite methods to identify likely pulmonary pathogens, we could not adjudicate the pathogenicity of each microbe or contribution to each patient’s pulmonary disease. Sixth, clinical microbiologic testing of BAL varied across hospitals and was not standardized. Seventh, while our work implicates pulmonary microbial depletion in the pathobiology of post-HCT lung disease, we cannot prove causality with correlative human studies, and cannot account for effects from other microbiomes such as the intestinal microbiome on lung health.^84,85^

In summary, we present the largest investigation to date of the pulmonary microbiome and transcriptome in pediatric HCT patients. We identified four unique BAL clusters that combine microbiome and lung gene expression signatures. The worst outcomes were observed for those with commensal microbe depletion, viral or fungal enrichment, lymphocyte activation, and fibroproliferation. Overall, these findings represent a step forward in understanding lung disease biology in HCT patients and may be used to improve patient subtyping in preparation for a future biology-targeted clinical trial.

## METHODS

### Patients

The derivation cohort was enrolled through the Pediatric Transplantation and Cell Therapy Consortium (PTCTC, NCT02926612) and the validation cohort was collected at the University Medical Center in Utrecht, The Netherlands. Participating pediatric centers screened all patients with a history of allogeneic (both cohorts) or autologous (PTCTC cohort only) HCT preparing to undergo clinically-indicated bronchoscopic BAL for diagnostic assessment of pulmonary disease. Patients or their guardians were approached prospectively for consent under local IRB approval at each site and permission was obtained to collect leftover BAL fluid. Patients were excluded if there was a limitation of care such as do not resuscitate at the time of BAL.

### BAL specimen collection

Bronchoscopy and BAL were performed at the discretion of the treating team using local institutional protocols. All BAL were obtained by pediatric pulmonologists trained in fiberoptic bronchoscopy with anesthesia provided by anesthesiologists or critical care physicians. Lavage protocol was not dictated by the study but typically involved 3-6 aliquots of 10mL sterile saline inserted into diseased areas of the lung as determined by preceding chest imaging or physical exam.^86^ Percent of lavage returned was not routinely documented and lavage aliquots were typically pooled by the clinical team immediately after collection.^87,88^ After aliquoting for clinical testing, excess lavage was placed immediately on dry ice, stored at -70°, shipped to UCSF, and stored at -70°C until processing.

### Clinical protocols and data collection

Clinical microbiologic testing was determined by the treating team and typically included culture for bacteria, fungus, and AFB; multiplex PCR for respiratory viruses; galactomannan antigen; and cytology for PCP. Additional molecular diagnostics such as PCR for atypical bacteria or fungi were used at the discretion of the site. After BAL, supportive care protocols were determined by the treating team; all patients were enrolled at centers with pediatric intensive care units. Patient demographics, medical history, and transplant-specific data were documented by trained study coordinators at each site. The most recent ANC and ALC measured clinically prior to BAL were documented. Results of clinical microbiologic testing on BAL were documented and not considered complete until 4 weeks after collection. For the PTCTC cohort, all doses of antimicrobials administered in the 7 days prior to BAL were documented. Antibiotic exposure score (AES) was calculated by summing days of exposure to each antibacterial agent weighted with an agent-specific broadness score ranging from 4 to 49.75 (e.g.: ampicillin 13.50, meropenem 41.50).^38^ Daily dosages were not collected. The number of anti-anaerobe days were calculated as the sum of preceding exposure to each of the following: Amoxicillin/clavulanic acid, Ampicillin/sulbactam, Pipercillin/tazobactam, Meropenem, Ertapenem, Imipenem, Levofloxacin, Clindamycin, Doxycycline, Tigecycline, or Metronidazole. Patients were followed until hospital discharge (PTCTC) or until at least one year post-BAL (Utrecht) with no loss to follow-up.

### BAL RNA Extraction

Samples were used on the first or second thaw. Samples underwent a previously described RNA extraction protocol optimized for BAL fluid.^11^ 200 µL of BAL was combined with 200 µL DNA/RNA Shield (Zymo) and 0.5mm glass bashing beads (Omni) for 5 cycles of 25 seconds bashing at 30Hz, with 60 seconds of rest on ice between each cycle (TissueLyser II, Qiagen). Subsequently, samples were centrifuged for 10 minutes at 4°C and the supernatant was used for column-based RNA extraction with DNase treatment according to the manufacturer’s recommendations (Zymo ZR-Duet DNA/RNA MiniPrep Kit). Resultant RNA was eluted in 5 µL sterile water and stored at -70°C until sequencing library preparation.

### BAL RNA Sequencing

Samples underwent a previously described sequencing library preparation protocol optimized for BAL fluid.^19^ First, BAL RNA was dehydrated at 40°C for 25 minutes in a 384 well plate (GeneVac E-Z2). Second, sequencing libraries were prepared using miniaturized protocols adapted from the New England Biolabs Ultra II RNA Library Prep Kit (dx.doi.org/10.17504/protocols.io.tcaeise). Reagents were dispensed using the Echo 525 (Labcyte) and underwent Ampure-XP bead cleaning on a Beckman Coulter Biomek NX^P^ instrument. Libraries underwent 19 cycles of polymerase chain reaction (PCR) amplification, size selection to a target 300 to 700 nucleotides (nt), and were pooled to facilitate approximately even depth of sequencing. Twenty-five picograms (pg) of External RNA Controls Consortium (ERCC) pooled standards were spiked-in to each sample after RNA extraction and before library preparation to serve as internal positive controls (Thermo Fisher Scientific Cat. No 4456740). In addition, to identify contamination in laboratory reagents and the laboratory environment, each batch contained 2 samples of 200 µL sterile water and 6-8 samples of 200 µL HeLa cells taken from a laboratory stock and processed identically to patient samples, in order to account for laboratory- and reagent-introduced contamination. These samples were processed at the same time as the patient BAL samples in order to use the same lot of reagents and minimize batch effect on control samples. Samples were processed and sequenced in 4 batches. Samples were pooled across lanes of an Illumina NovaSeq 6000 instrument and sequenced to a target depth of 40 million read-pairs with sequencing read length of 125 nt.

### Sequencing file processing

All sequencing files were processed using the CZID pipeline v7.1 (https://github.com/chanzuckerberg/czid-web).^18^ Briefly, .fastq files underwent a first round of human read subtraction (*STAR* to hg38) followed by Illumina adaptor removal (*Trimmomatic*), quality filtering (*PriceSeq*), duplicate read removal (*CD-HIT-DUP*), and LZW complexity filtering. Next, sequencing files underwent a second more stringent round of human read subtraction (*Bowtie2*) followed by a third round of human read subtraction (*STAR*), subsampling to 1 million fragments, and a fourth and final round of human read subtraction (*GSNAP*). Human gene counts were produced using the CZID pipeline with alignment to hg38 as described above. 60,590 total genes were detected across all samples (median 44,063, IQR 31,553-52,129), and were subset for 19,032 protein coding genes (median 18,259 genes per sample, IQR 16,988-18,871) and used in analyses below. Resultant human-subtracted sequencing files were then used in two ways for microbiome characterization:

***(1) Microbial taxonomic alignment:*** Human-subtracted sequencing files underwent alignment to the NCBI nt/nr database using GSNAP with minimum alignment length >36. Quality metrics for the sequencing run including percent of reads that passed the PriceSeq filter step and percent of reads that passed all steps were examined and samples with poor sequencing quality were re-sequenced. Taxa counts were generated with associated metrics of percent identity, contig length, and e-value to the nearest NCBI hit. To reduce spurious associations due to ambiguous alignments, taxa were excluded if they (1) aligned to archaea or uncultured microbes, (2) had ≤6 total reads, (3) had <100 nt alignment length, or (4) had <80%, <90%, or <95% nt percent identity for viruses, eukaryotes, and bacteria, respectively. In addition, samples with low biomass (<100pg) were further filtered to keep only taxa with ≥10 transcripts forming a contig of ≥250 nt with ≥80% percent identity to the nearest NCBI hit.
***(2) Microbial functional alignment:*** Human-subtracted sequencing files were processed using FMAP v.0.15^89^ in order to profile the metabolic pathways present in each sample. FMAP_mapping.pl was paired with diamond v.0.9.24^90^ and FMAP_quantification.pl were used with default settings to identify and quantify associated proteins in the UniRef90 database.^91,92^ The gene assignments were regrouped by KEGG descriptors ^93,94^ and their annotation was summarized at levels 1 to 3.

### Microbial quantification and contamination

Low biomass samples are susceptible to contamination.^25,27^ We previously showed that a positive control spike-in to each sample can be used to back-calculate the original RNA mass of the sample by solving the linear proportionality equation (total sample reads / total sample mass) ≈ (ERCC reads / ERCC mass), where sample reads and ERCC reads were detected by the above protocol and ERCC input was standardized as 25 pg.^20^ The calculated sample mass was then reduced by 25 pg (the ERCC input) to equal the original sample mass before ERCC addition. Since the input RNA mass of the water controls was determined to be about (5 pg presumably reflecting 5 pg of sequenceable contamination), we discarded samples whose total input mass was below 10 pg, as we were unable to reliably differentiate between contamination and true constituents. Since low biomass samples will preferentially amplify contaminants, we then used the ERCC spike-in to transform reads into estimated mass, allowing analysis of both fractional and absolute microbiome properties. Since each BAL microbiome consists of contributions from the patient and externally introduced contaminants, we then calculated the unique contamination profile of the water and HeLa samples for each sequencing batch, and subtracted the mean + 2SD of each contaminant taxa from the patient samples processed in the respective batch. Mass-transformed and contamination-adjusted values were used for downstream analysis.

### Statistical Analysis

***(1) Unsupervised Clustering Analysis:*** Since microbiome data can be described using taxonomy, functional annotation, or summary measures, we used the Multi-Omics Factor Analysis to reduce dimensionality and identify a core set of factors.^95^ This approach accommodates different data structures and distributions and is tolerant of collinearity. Data were filtered to include phyla, genera, species, and KEGG pathways present in >15% of samples, underwent variance stabilizing transformation (*vst, DESeq2*), and were combined with aggregate metrics of total microbial mass, Simpson’s and Shannon’s alpha diversity (*vegan*), and richness, which was defined as number of species detected at a threshold of ≥ 1 pg.^96,97^ MOFA was used to identify 15 core latent factors that together explained the most variance in the data structure. The matrix of latent factor values then underwent uniform manifold approximation mapping (*umap*) and BAL clusters were identified using hierarchical clustering of euclidean distances (*eclust*, *factoextra*). The ideal number of clusters was determined to be four using the silhouette, elbow, and gap-statistic plots.
***(2) Clinical characteristics:*** Kaplan Meier survival analysis was used to plot in-hospital mortality by BAL cluster and survival curves were compared using the log-rank test of equality (*survival*). Differences in clinical traits across clusters (eg: antimicrobial exposure score, absolute neutrophil count) were tested using the non-parametric Kruskal-Wallis (*kruskaltests*) and Dunn’s tests (*dunn.test*) or Chi-squared test as appropriate.
***(3) Microbiome comparisons:*** Differences in microbial taxa, KEGG pathways, richness, and diversity across the 4 BAL clusters were tested using the non-parametric Kruskal-Wallis (*kruskaltests*) and Dunn’s tests (*dunn.test*) with Benjamini-Hochberg correction for multiple hypothesis testing. Differences in microbial taxa and KEGG pathways were also tested using negative binomial generalized linear models, which account for both microbiome composition and size by inclusion of taxa-specific dispersion factors (*edgeR*).^98^ Associations between microbial taxa and clinical variables (e.g.: antimicrobial exposure score, in-hospital mortality) were tested using *edgeR*. Data were visualized with heatmaps showing cluster means for each variable (*pheatmap*) with individual comparisons shown using box-whisker plots (*ggplot*). Causal mediation was used to test whether the association between antimicrobial exposure and mortality was mediated by an antibiotic-induced reduction in certain BAL microbes (*mediation*).^99,100^ Using the latent structural equation framework, we fit (1) poisson models for the association between preceding AES and BAL quantity of a certain microbe, and (2) logistic regression models for the association between BAL quantity of a given microbe and outcome, independent of AES. Mediation was tested using 1,000 simulations with bootstrapped confidence intervals and direct and indirect effects were plotted (**eFigure 7**).
***(4) Pathogen identification:*** Taxa considered as potential respiratory pathogens were adapted from the CZID Pathogen List (https://czid.org/pathogen_list) with modifications for immunocompromised patients and pathogens specific to the respiratory system. The final list of taxa considered is detailed in **Data File 4**. We did not include avirulent viruses, such as TTV, or bacterial commensals that are infrequently a cause of pulmonary disease, such as Prevotella species, coagulase-negative Staphylococci, non-diphtheria Corynebacterium, and viridans group Streptococci, although these have at times been implicated in pulmonary disease in immunocompromised patients. To identify potentially pathogenic viruses, we applied a threshold of viral detection at any level above background (after applying the quality and contamination filters described above). This presence/absence approach was selected to mirror the approach used in clinical respiratory viral panels, which typically dichotomizes any level of detection as present/absent. To identify potentially pathogenic bacteria, we applied a threshold of detection with mass ≥10pg, bacterial dominance ≥20%, and Z-score ≥ +2, where Z-score was calculated as the number of standard deviations above the mean of the log_10_-transformed mass values for each microbe in the cohort. Requiring a minimum mass, dominance, and z-score was based on the historical framework that bacterial infections occur when microbes are present at high mass that is greater than other microbes and greater than in other (non-infected) patients, although this may not be true in all instances. Cutoff values were empirically selected after analysis of data distributions and could be exchanged for other cutoffs in order to alter the balance between sensitivity and specificity of calls. Finally, to identify potentially pathogenic fungi, we applied a threshold of detection with mass ≥10pg and Z-score ≥ +2. We did not apply a microbiome dominance cutoff for fungal pathogens since the relationship between organisms in the pulmonary mycobiome is less well understood.
***(5) Gene expression:*** Only genes present in >25% of samples were used for differential gene expression. To identify individual differentially expressed genes, we used a 4-way ANOVA-like approach with negative binomial generalized linear models (*edgeR*). Select differentially expressed genes identified at a threshold FDR ≤0.05 were visualized with box-whisker plots of variance stabilization-transformed counts. To compute gene set enrichment scores, we used non-parametric gene set variation analysis with Poisson distributions (*gsva*) and the Reactome set of n=1,554 gene sets.^101,102^ Differences in enrichment scores across the BAL clusters were compared using Kruskal-Wallis (*kruskaltests*) and Dunn’s tests (*dunn.test*) and gene sets with significant differences were visualized using dot plots of the mean expression scores (*pheatmap*). Next, cell types contributing to bulk seq expression were imputed using CIBERSORTx (Docker version), which employs a user-defined reference single-cell atlas to identify cell-type specific transcript ratios and impute cell fractions (we selected the *Travaglini et al* lung cell atlas).^42–44^ Cell-type specific gene expression was imputed using CIBERSORTx high resolution mode, which utilizes previously created cell fractions to impute cell-type specific expression. Finally, lymphocyte receptor repertoires were imputed using Imrep (Linux install), which identifies CDR3 alignments from within bulk gene expression data.^103,104^
***(6) Classification and validation:*** Since cluster assignments cannot be directly applied to an external dataset, a classification tool is required to predict cluster assignments. We trained a random forest of n=10,000 trees using microbiome taxonomy and lung gene expression datasets as input, and 2x weighting of clusters 3 and 4 given the BAL cluster imbalance (*randomforestSRC*).^105,106^ Ideal forest parameters determined using *tune* were similar to default settings and thus default settings were used for all other parameters (eg: *mtry*, *nodesize*, etc). Forest accuracy was determined using out of bag AUC and a confusion matrix. Variable importance was determined using permutation VIMP (Breiman-Cutler importance) by permuting OOB cases (*vimp*). To validate the classifier, the random forest classifier was applied to microbiome taxonomy and lung gene expression data from the n=57 Utrecht BALs and 1-year post-BAL non-relapse mortality rates were compared according to predicted BAL cluster type using Kaplan-Meier survival curves with the log-rank test.

## DATA AVAILABILITY

Sequencing files are posted on dbGaP: https://www.ncbi.nlm.nih.gov/projects/gap/cgi-bin/study.cgi?study_id=phs001684.v2.p1

## AUTHOR CONTRIBUTIONS

M.S.Z., C.C.D., G.A.Y., M.A.P., and J.L.D. contributed to the conception and design of the work. All authors contributed to the acquisition of data. M.S.Z., C.C.D., M.Y.M., G.R., M.R.S., E.M.P., H.K., I.S., L.N.S., and J.L.D. contributed to the analysis of data. All authors contributed to the drafting and revision of the manuscript.

## ACKNOWLEDGEMENTS

M.S.Z. received research funding from NHLBI K23HL146936, NICHD K12HD000850, the American Thoracic Society, the Pediatric Transplantation and Cell Therapy Foundation, and the National Marrow Donor Program Amy Strelzer Manasevit Grant. M.Y.M received research funding from NCI F31CA271571. H.A-A. received grant funding from the Gateway Foundation and St. Baldrick’s Foundation. J.S.K. and J.J.B. received research funding from NCI P30CA008748.

M.A.P. received research funding from NCI P30CA040214. L.N.S. received research funding from NIGMS R21GM147800, NCI R37CA244775, and NCI U2CCA271890. J.L.D. received research funding from the Chan Zuckerberg Biohub. Additional funding for the study was provided by NHLBI UG1HL069254 and a Johnny Crisstopher Children’s Charitable Foundation St. Baldrick’s Consortium Grant.

