## Supplemental Data for "Pulmonary microbiome and transcriptome signatures reveal distinct pathobiologic states associated with mortality in two cohorts of pediatric stem cell transplant patients"

**Supplement**

**Contents:**

**Tables**


1. eTable 1: PTCTC Site Principal Investigators & Support Staff
2. eTable 2: Clinical Background, Stratified by Cluster
3. eTable 3: Illness Severity and Clinical Outcomes, Stratified by Cluster
4. eTable 4: In-Hospital Mortality by Cluster, Subset for Patients ≤100 Days Post-HCT
5. eTable 5: Multivariable Cox Model for In-Hospital Mortality According to BAL Cluster
6. eTable 6. Diagnostic Agreement Table for Potential Pathogens Detected in BAL by Clinical and Metagenomic Approaches
7. eTable 7: Torquetenovirus Detection, Stratified by Cluster
8. eTable 8: Hospital test vs Metagenomic Sequencing
9. eTable 9: In-Hospital Mortality Stratified by Hospital-Based vs Metagenomics Results
10. eTable 10: Cluster transitions from 1st to 2nd+ BAL
11. eTable 11: Cluster prevalence according to 1st vs 2nd+ BAL
12. eTable 12: Random Forest Classifier
13. eTable 13: Validation Cohort Characteristics
14. eTable 14: Validation Cohort Cluster Assignments


**Figures**


1. eFigure 1: HCT Day and Immune Counts at Time of BAL
2. eFigure 2: MOFA Factor Correlation
3. eFigure 3: Percent of Variance Explained by MOFA Factors
4. eFigure 4: Selection of Optimal Number of Clusters
5. eFigure 5: Representative BALs from Each Cluster
6. eFigure 6: BAL Microbial Expression of KEGG for Carbohydrate, Energy, and Lipid metabolism pathways.
7. eFigure 7: BAL Microbial Expression of KEGG Glycan Biosynthesis Pathways
8. eFigure 8: In-Hospital Survival Stratified by Metagenomics Results
9. eFigure 9: In-Hospital Survival Stratified by Antibacterial Exposure Score
10. eFigure 10: Causal Mediation of Effect of Antibiotic Exposure on Outcome
11. eFigure 11: Imputed BAL Cell Fractions
12. eFigure 12: Imputed Monocyte/Macrophage-Specific Gene Expression
13. eFigure 13: Imputed Lymphocyte-Specific Gene Expression
14. eFigure 14: T-Cell Receptor Alpha Reads
15. eFigure 15: BAL Cluster Transitions
16. eFigure 16: Validation Cohort Survival Curves

**Data Files**

1. Kruskal-Wallis testing identifying BAL taxa associated with cluster assignment
2. NB-GLM identifying BAL taxa associated with in-hospital mortality
3. Kruskal-Wallis testing identifying BAL KEGG pathways associated with cluster assignment
4. List of microbes considered clinically-relevant pulmonary pathogens
5. Patient-level comparison of hospital infectious tests to MNGS pathogen results
6. NB-GLM identifying BAL taxa associated with antibacterial exposure score (AES)
7. BAL taxa mediating the association between AES and in-hospital mortality
8. BAL taxa mediating the association between anti-anaerobic score and in-hospital mortality
9. NB-GLM identifying BAL human transcripts associated with cluster assignment
10. Kruskal-Wallis testing identifying BAL Reactome pathways associated with cluster
11. NB-GLM identifying BAL human transcripts associated with in-hospital mortality
12. Variable importance for variables used in random forest BAL classifier

**Raw Sequencing Files**

1. dbGaP https://www.ncbi.nlm.nih.gov/projects/gap/cgi-bin/study.cgi?study_id=phs001684.v2.p1

**eTable 1: PTCTC Site Principal Investigators & Support Staff**

| Site | Site Name | Site PI | Specialty | Research Coordinators, Support Staff |
|---|---|---|---|---|
| AUS | Children's Hospital at Westmead | Peter J. Shaw | BMT | Jun Cai |
| AZ | Phoenix Children's Hospital | Erin M. Kreml | ICU | Brian Burrows, Jessica Twyford |
| CA | Children’s Hospital Los Angeles | Hisham Abdel-Azim | BMT | Anju Nair, Sandy Gutierrez |
| CA | UCLA | Theodore B. Moore | BMT | LaMarr Taylor Smith, Andres Vargas |
| CA | UCSF-San Francisco | Matt Zinter | ICU | Kevin Magruder, Anne McKenzie |
| CA | UCSF-Oakland | Nahal Lalefar | BMT | Ad hoc |
| CAN | Cancer Care Manitoba | Geoffrey D.E. Cuvelier | BMT | Laura Keuler, Erin Richards |
| CO | Children's Hospital Colorado | Amy K. Keating | BMT | Garrett Donegan, Joanna Wozniak, Steven Kyker |
| DC | Children's National | Benjamin Hanisch | ID | Alexa Yarish |
| FL | Nicklaus Children's | Kamar Godder | BMT | Guido Elias, Kristofer Rosales |
| FL | University of Florida, Gainesville | Paul Castillo | BMT | Giselle Moore-Higgs, Beate Greer, Joshua Terrell |
| GA | Emory | Muna Qayed | BMT | Andrea Peters, Ashley Griffin |
| IL | Lurie Children's | Sonali Chaudhury | BMT | Olga Jonas, Dian’Ella Ramsey |
| IN | Riley Children's | Courtney M. Rowan | ICU | Kirsten Ramberg, Jill Mazurczyk, Andrea Hudgins |
| MA | Boston Children's | Christine N. Duncan | BMT | Lauren Leonard, Sophie Silverstein, Miranda Fry |
| MI | Helen DeVos Children's | Aly Abdel-Mageed | BMT | Jennifer Hanenburg |
| MN | University of Minnesota | Janet R. Hume | ICU | Lexie Goertzen |
| MN | Mayo Clinic | Shakila P. Khan | BMT | Julia Byrne, Becky Winslow-Rain |
| MS | Children's Hospital Mississippi | Dereck B. Davis | BMT | Sarah Elkin, Katie Williams, Michelle Kimble |
| NC | Duke University | Paul L. Martin | BMT | Lucy Harris, Linda Brown |
| NJ | Hackensack | Shira J. Gertz | ICU | Elana Smilow, Gina Dovi, Jeanette Haugh |
| NY | Columbia | Prakash Satwani | BMT | Aaleen Cox, Chez Brivett, Brian Volonte |
| NY | Memorial Sloan Kettering/Cornell | James S. Killinger | ICU | Jennifer Sollitto, Jennifer Direnzo |
| OH | Cleveland Clinic | Rabi Hanna | BMT | Rawan Nawabit, Alexis Smith |
| OH | Nationwide | Jeffrey J. Auletta | BMT | Clelie Peck, Kelly Ortman, Mary Scholz |
| PA | Children’s Hospital of Philadelphia | Julie C. Fitzgerald | ICU | Jenny Bush, Mary Diliberto, Martha Sisko |
| PA | Pittsburg | Jessie L. Barnum | BMT | Olga Greg |
| SC | Medical University South Carolina | Michelle P. Hudspeth | BMT | Jared Hortman |
| TN | St. Jude | Caitlin Hurley | ICU | Ad hoc |
| TX | MD Anderson | Kris M. Mahadeo | BMT | LaTarsha Williams, So Jung Hong |
| TX | Methodist San Antonio | Troy C. Quigg | BMT | Candace Taylor, Marisa Palacios |
| WI | Medical College of Wisconsin | Rachel Phelan | BMT | Adam Fiebelkorn, Melissa Schussman |

**eTable 2: Clinical Background, Stratified by Cluster**

| **Demographics (n=229 patients)** | **Cluster 1 (n=101)** | **Cluster 2 (n=59)** | **Cluster 3 (n=43)** | **Cluster 4 (n=26)** | **Sig.** |
|---|---|---|---|---|---|
| Age (median years, IQR) | 11.5 (4.5-17.4) | 11.0 (5.4-15.1) | 10.5 (5.1-14.5) | 10.6 (4.2-16.6) | P=0.924 |
| Sex (male) | 64 (64%) | 40 (68%) | 18 (42%) | 11 (42%) | **P=0.013** |
| Race | | | | | P=0.071 |
| - White, not Latinx | 68 (67%) | 31 (53%) | 27 (63%) | 14 (54%) | |
| - Black | 9 (9%) | 9 (15%) | 8 (19%) | 3 (12%) | |
| - Other/multiple | 11 (11%) | 4 (7%) | 4 (9%) | 7 (27%) | |
| - Asian/PI | 10 (10%) | 11 (19%) | 3 (7%) | 1 (4%) | |
| - Native American | 1 (1%) | 0 (0%) | 0 (0%) | 1 (4%) | |
| - Unknown | 2 (2%) | 4 (7%) | 1 (2%) | 0 (0%) | |
| Ethnicity - Latino/Hispanic | 25 (25%) | 11 (19%) | 12 (28%) | 11 (42%) | P=0.142 |
| **Medical History (n=229 patients)** | | | | | |
| Disease | | | | | |
| - Leukemia[a] | 55 (54%) | 35 (59%) | 21 (49%) | 14 (54%) | P=0.599 |
| - Inborn errors of immunity[b] | 19 (19%) | 9 (15%) | 8 (19%) | 4 (15%) | |
| - Non-malignant hematologic[c] | 7 (7%) | 7 (12%) | 9 (21%) | 4 (15%) | |
| - Solid tumor[d] | 7 (7%) | 2 (3%) | 3 (7%) | 2 (8%) | |
| - Lymphoma[e] | 7 (7%) | 3 (5%) | 2 (5%) | 0 (0%) | |
| - Inborn errors of metabolism[f] | 6 (6%) | 3 (5%) | 0 (0%) | 2 (8%) | |
| HCT Type | | | | | |
| - Allogeneic | 92 (91%) | 57 (97%) | 40 (93%) | 24 (92%) | P=0.520 |
| - Bone marrow | - 38 (41%) | - 27 (47%) | - 20 (50%) | - 7 (29%) | |
| - Peripheral blood | - 36 (39%) | - 24 (42%) | - 15 (38%) | - 13 (54%) | |
| - Umbilical cord blood (UCB) | - 18 (20%) | - 6 (11%) | - 5 (13%) | - 4 (17%) | |
| - Autologous | 9 (9%) | 2 (3%) | 3 (7%) | 2 (8%) | |
| HLA match (allogeneic only) | | | | | P=0.950 |
| - Matched related donor | 20 (22%) | 13 (23%) | 7 (18%) | 4 (21%) | |
| - Matched unrelated donor (inc. 6/6 UCB) | 18 (20%) | 14 (25%) | 9 (23%) | 8 (33%) | |
| - Mismatched related donor (haplo) | 27 (29%) | 13 (23%) | 11 (28%) | 6 (25%) | |
| - Mismatched unrelated donor (inc. <6/6 UCB) | 27 (29%) | 17 (30%) | 13 (33%) | 5 (21%) | |
| Conditioning Agents Used[g] | | | | | |
| - Backbone agent | | | | | |
| - Busulfan | 33 (33%) | 24 (41%) | 16 (37%) | 13 (50%) | P=0.393 |
| - Melphalan | 28 (28%) | 16 (27%) | 11 (26%) | 8 (31%) | P=0.973 |
| - Total body irradiation (TBI) | 31 (31%) | 14 (24%) | 9 (21%) | 5 (19%) | P=0.472 |
| - Other alkylating agent | | | | | |
| - Cyclophosphamide | 43 (43%) | 21 (36%) | 16 (37%) | 11 (42%) | P=0.814 |
| - Thiotepa | 29 (29%) | 17 (29%) | 14 (33%) | 6 (23%) | P=0.870 |
| - Antimetabolite | | | | | |
| - Clofarabine | 6 (6%) | 4 (7%) | 2 (5%) | 3 (12%) | P=0.711 |
| - Fludarabine | 61 (60%) | 41 (69%) | 29 (67%) | 15 (58%) | P=0.572 |
| - Serotherapy (ATG or Alemtuzumab) | 51 (51%) | 33 (56%) | 21 (49%) | 14 (54%) | P=0.881 |

**Legend:** Characteristics compared using Kruskal-Wallis test or Chi-squared test, as appropriate.

**eTable 3: Clinical Presentation and Outcomes, Stratified by Cluster**

| Characteristics at Enrollment (n=278 BALs) | Cluster 1 N=127 | Cluster 2 N=74 | Cluster 3 N=45 | Cluster 4 N=32 | Sig |
|---|---|---|---|---|---|
| Days from HCT to BAL (n, %) | 105 (28-309) | 218 (50-476) | 82 (37-146) | 121 (47-351) | **P=0.016** |
| Days from Symptoms to BAL[a] (n, %) | 6 (2-14) | 11 (3-30) | 8 (2-22) | 9 (3-28) | P=0.190 |
| Clinical Presentation Symptoms (n, %) | | | | | |
| - Lower respiratory symptoms) | 104 (82%) | 71 (96%) | 43 (96%) | 31 (97%) | **P=0.002** |
| - Hypoxia ≤96% | 93 (73%) | 46 (62%) | 38 (84%) | 25 (78%) | P=0.051 |
| - Abnormal chest x-ray | 78/88 (89%) | 43/56 (77%) | 32/39 (82%) | 21/24 (88%) | P=0.271 |
| - Abnormal chest CT | 102/107 (95%) | 51/51 (100%) | 31/34 (91%) | 25/26 (96%) | P=0.243 |
| - Worse PFTs | 10 (8%) | 2 (3%) | 1 (2%) | 3 (9%) | P=0.248 |
| Oxygen Required Prior to BAL | | | | | |
| - No | 79 (62%) | 47 (64%) | 16 (36%) | 14 (44%) | **P=0.004** |
| - Yes | 48 (38%) | 27 (36%) | 29 (64%) | 18 (56%) | |
| Oxygen Type Prior to BAL | | | | | |
| - None | 79 (62%) | 47 (64%) | 16 (36%) | 14 (44%) | **P=0.019** |
| - Nasal cannula or face mask | 18 (14%) | 10 (14%) | 9 (20%) | 4 (13%) | |
| - HFNC, NIV, or Intubation | 30 (24%) | 17 (23%) | 20 (44%) | 14 (44%) | |
| Comorbidities at time of BAL (n, %) | | | | | |
| - Engraftment syndrome | 10 (8%) | 3 (4%) | 2 (4%) | 0 (0%) | P=0.296 |
| - GVHD active at time of BAL | 29 (25%) | 28 (39%) | 11 (27%) | 15 (48%) | **P=0.040** |
| - GVHD ever preceding BAL | 46 (40%) | 42 (58%) | 18 (44%) | 20 (65%) | **P=0.019** |
| - Heart failure or reduced function | 4 (3%) | 2 (3%) | 3 (7%) | 2 (6%) | P=0.607 |
| - Kidney injury | 16 (13%) | 7 (9%) | 14 (31%) | 10 (31%) | **P=0.001** |
| - Pericardial effusion | 13 (10%) | 3 (4%) | 5 (11%) | 4 (12%) | P=0.366 |
| - Pulmonary hemorrhage/hemoptysis | 14 (11%) | 2 (3%) | 3 (7%) | 4 (13%) | P=0.158 |
| - Sepsis | 16 (13%) | 11 (15%) | 7 (16%) | 3 (9%) | P=0.841 |
| - TA-TMA | 10 (8%) | 5 (5%) | 3 (7%) | 5 (16%) | P=0.341 |
| - VOD/SOS | 11 (9%) | 3 (4%) | 6 (13%) | 4 (13%) | P=0.280 |
| Immunologic Function Prior to BAL | | | | | |
| - WBC (median. IQR) | 4,230 (2,300-7,300) | 5,700 (2,500-12.5k) | 4,400 (1,500-7,790) | 4,685 (3,400-8,750) | P=0.098 |
| - ANC (median, IQR) | 2,774 (1,233-4,581) | 4,059 (1,960-7,155) | 2,490 (940-5,125) | 3,344 (2,007-5,640) | **P=0.029** |
| - ANC <0.5 $x10^9$/L (n, %) | 19 (15%) | 5 (7%) | 9 (20%) | 1 (3%) | **P=0.047** |
| - ALC (median, IQR) | 423 (179-994) | 414 (131-1,102) | 417 (155-1,106) | 405 (196-1,187) | P=0.997 |
| - ALC <0.2 $x10^9$/L (n, %) | 34 (27%) | 21 (28%) | 13 (29%) | 8 (25%) | P=0.977 |
| **Outcomes** | | | | | |
| **Intensive Care After BAL** (yes/no) | 64 (50%) | 29 (39%) | 34 (76%) | 20 (62%) | **P=0.001** |
| **Mechanical Ventilation After BAL** | | | | | |
| - ≥ 24 hours | 49 (39%) | 23 (31%) | 26 (58%) | 16 (50%) | **P=0.022** |
| - ≥ 7 days | 33 (26%) | 16 (22%) | 23 (51%) | 14 (44%) | **P=0.001** |
| **In-Hospital Mortality (n=229 patients)** | N=101 | N=59 | N=43 | N=26 | |
| - By 14 days | 6 ( 6%) | 2 ( 3%) | 3 ( 7%) | 6 (23%) | **P=0.012** |
| - By 28 days | 9 ( 9%) | 4 ( 7%) | 7 (16%) | 7 (27%) | **P=0.032** |
| - All In-hospital | 14 (14%) | 8 (14%) | 14 (33%) | 9 (35%) | **P=0.008** |
| **In-Hospital Mortality if requiring oxygen prior to BAL (n=103 patients)** | N=37 | N=23 | N=28 | N=15 | |
| - By 14 days | 2 ( 5%) | 2 ( 9%) | 3 (11%) | 6 (40%) | **P=0.006** |
| - By 28 days | 4 (11%) | 4 (17%) | 7 (25%) | 7 (47%) | **P=0.035** |
| - All In-hospital | 8 (22%) | 7 (30%) | 14 (50%) | 9 (60%) | **P=0.022** |

**Legend:** Since some patients had multiple BALs, in-hospital mortality was calculating using the most recent BAL. Characteristics compared using Kruskal-Wallis test or Chi-squared test, as appropriate.

**eTable 4: In-Hospital Mortality by Cluster, Subset for Patients ≤100 Days Post-HCT**

| | **Cluster 1 N=51** | **Cluster 2 N=24** | **Cluster 3 N=25** | **Cluster 4 N=12** | **Significance** |
|---|---|---|---|---|---|
| **In-Hospital Mortality** | | | | | |
| - **Yes** | 6 (12%) | 7 (29%) | 10 (40%) | 6 (50%) | P=0.019 |
| - **No** | 45 (88%) | 17 (71%) | 15 (60%) | 6 (50%) | |

**Legend:** Only the most recent BAL for each patient was used. Significance tested with Chi-squared test.

**eTable 5: Multivariable Cox Model for In-Hospital Mortality**

| | **Hazard Ratio (95% CI)** | **Significance** |
|---|---|---|
| **BAL Cluster (relative to Cluster 1)** | | |
| - **Cluster 2** | 1.19 (0.44-3.20) | P=0.734 |
| - **Cluster 3** | 2.63 (1.10-6.31) | **P=0.030** |
| - **Cluster 4** | 3.43 (1.34-8.80) | **P=0.010** |
| **Age (years)** | 0.99 (0.94-1.05) | P=0.826 |
| **Biologic Sex (male reference)** | 0.64 (0.33-1.25) | P=0.191 |
| **ANC** | 1.04 (0.98-1.10) | P=0.268 |
| **ALC** | 0.35 (0.17-0.75) | **P=0.007** |
| **GVHD (yes/no)** | 1.17 (0.59-2.33) | P=0.658 |

**Legend:** Cox multivariable model for in-hospital mortality.

**eTable 6. Diagnostic Agreement Table for Potential Pathogens Detected in BAL by Clinical and Metagenomic Approaches**

| | C | C+M | M | | C | C+M | M | | C | C+M | M |
|---|---|---|---|---|---|---|---|---|---|---|---|
| **Community Viruses** | | | | **Cultivable Bacteria** | | | | **Fungi** | | | |
| Adenovirus | 4 | 4 | 7 | Achromobacter | 0 | 1 | 0 | Alternaria | 0 | 0 | 4 |
| Coronavirus | 1 | 3 | 5 | Bacillus | 0 | 0 | 1 | Aspergillus [a] | 13 | 2 | 16 |
| Influenza virus | 1 | 1 | 3 | Citrobacter | 0 | 0 | 1 | Candida | 3 | 2 | 19 |
| Metapneumovirus | 0 | 1 | 1 | Escherchia | 1 | 3 | 8 | Cladosporium | 0 | 0 | 3 |
| Parainfluenza virus | 1 | 3 | 3 | Enterococcus | 2 | 1 | 2 | Cryptococcus | 0 | 0 | 1 |
| Rhinovirus | 5 | 27 | 20 | Haemophilus | 3 | 8 | 3 | Exophiala | 0 | 0 | 4 |
| RSV | 0 | 2 | 2 | Klebsiella | 2 | 1 | 4 | Exserohilum | 0 | 0 | 8 |
| | | | | Moraxella | 1 | 3 | 2 | Fusarium | 1 | 0 | 17 |
| **Herpesviruses** | | | | Pseudomonas | 1 | 9 | 2 | Mucor | 0 | 1 | 6 |
| Cytomegalovirus | 9 | 14 | 9 | Salmonella | 0 | 0 | 1 | Pneumocystis | 1 | 2 | 6 |
| Epstein-Bar Virus | 3 | 0 | 8 | Staphylococcus | 3 | 3 | 1 | Saccharomyces | 0 | 1 | 13 |
| Herpes Simplex Virus 1 | 2 | 0 | 6 | Stenotrophomonas | 1 | 4 | 12 | | | | |
| Human Herpes Virus-6 | 5 | 4 | 13 | Streptococcus | 1 | 1 | 1 | **Parasites** | | | |
| Human Herpes Virus-7 | 1 | 0 | 3 | | | | | Acanthamoeba | 0 | 0 | 4 |
| Varicella Zoster Virus | 0 | 0 | 0 | **Atypical Bacteria** | | | | Toxoplasma | 0 | 0 | 4 |
| | | | | Chlamydia | 0 | 0 | 1 | | | | |
| **Other Viruses** | | | | Legionella | 0 | 0 | 0 | | | | |
| Bocavirus | 0 | 0 | 4 | Mycoplasma | 0 | 0 | 4 | | | | |
| BK polyoma | 0 | 0 | 1 | Ureaplasma | 0 | 0 | 1 | | | | |
| KI polyoma | 0 | 0 | 10 | | | | | | | | |
| LCMV | 0 | 0 | 1 | **Fastidious Bacteria** | | | | | | | |
| Parvovirus B19 | 0 | 0 | 1 | Actinomyces | 0 | 1 | 1 | **Total:** | 69 | 104 | 256 |
| Rubella | 0 | 0 | 1 | Mycobacteria | 2 | 0 | 0 | | | | |
| WU polyoma | 0 | 0 | 8 | Nocardia | 2 | 1 | 0 | | | | |

**Legend:** Potentially pathogenic microbes detected by clinical testing (C), metagenomic testing (M), or both clinical and metagenomic testing (C+M). Samples with multiple pathogens will contribute multiple entries in the table. Refer to **Data File 4** for list of potentially pathogenic taxa. Microbes not typically considered pulmonary pathogens, such as *S.epidermidis*, *P.melaninogenica*, and *R.mucilaginosa*, were not included. Refer to **Data File 5** for patient-level analysis. Refer to text for algorithm for discriminating potential pathogens from the background pulmonary microbiome. [a] While overlap between clinical and sequencing-based Aspergillus detection was low, of the n=13 Aspergillus detected clinically without NGS confirmation, all showed “one” or “rare” colonies on culture and only one was associated with a positive BAL galactomannan (0.589). Additionally, n=10 samples showed a positive BAL galactomannan and yet had no Aspergillus detected clinically or by sequencing with the above thresholds, suggesting a high rate of false-positivity for hospital-based testing.[28,29]

**eTable 7: Torquetenovirus Detection, Stratified by Cluster**

| All BALs (n=278) | Cluster 1 N=127 | Cluster 2 N=74 | Cluster 3 N=45 | Cluster 4 N=32 | Significance |
|---|---|---|---|---|---|
| **TTV Detected** | | | | | |
| - **Yes** | 10 (8%) | 23 (31%) | 12 (27%) | 10 (31%) | P<0.001 |
| - **No** | 117 (92%) | 51 (69%) | 33 (73%) | 22 (69%) | |

**Legend:** BAL detection of Alphatorquevirus, Betatorquevirus, or Gammatorquevirus RNA above background controls. Significance tested with Chi-squared test.

**eTable 8: Comparison of Hospital-Based vs Metagenomics Results**

| | Metagenomics Positive | Metagenomics Negative | McNemar's Test |
|---|---|---|---|
| **Any pathogen** | | | |
| - Hospital testing positive | 102 | 14 | P<0.001 |
| - Hospital testing negative | 91 | 71 | |
| **Any virus** | | | |
| - Hospital testing positive | 59 | 17 | P<0.001 |
| - Hospital testing negative | 51 | 151 | |
| **Any bacteria** | | | |
| - Hospital testing positive | 35 | 16 | P<0.001 |
| - Hospital testing negative | 42 | 185 | |
| **Any eukaryote** | | | |
| - Hospital testing positive | 10 | 15 | P<0.001 |
| - Hospital testing negative | 73 | 180 | |

**Legend:** Comparison of 2 diagnostic tests performed on same samples using McNemar's test

**eTable 9: In-Hospital Mortality Stratified by Hospital-Based vs Metagenomics Results**

| | Metagenomics Positive | Metagenomics Negative | Significance |
|---|---|---|---|
| **Any pathogen** | | | |
| - Hospital testing positive | 21/79 (27%) | 1/13 (8%) | P=0.190 |
| - Hospital testing negative | 15/80 (19%) | 8/57 (14%) | |

**Legend:** Significance assessed with the Chi-squared test.

**eTable 10: Cluster transitions from 1$^{st}$ to 2$^{nd}$+ BAL**

| | 2nd+ sample | | | |
|---|---|---|---|---|
| **1st sample** | **Cluster 1** | **Cluster 2** | **Cluster 3** | **Cluster 4** |
| **Cluster 1** | 9 | 6 | 2 | 9 |
| **Cluster 2** | 5 | 10 | 0 | 0 |
| **Cluster 3** | 0 | 0 | 0 | 2 |
| **Cluster 4** | 3 | 0 | 1 | 2 |

**Legend:** Raw data for BAL cluster transitions for each patient contributing ≥2 BALs to the cohort.

**eTable 11: Cluster prevalence according to 1$^{st}$ vs 2$^{nd}$+ BAL**

| | **1st BAL (n=229)** | **2nd+ BAL (n=49)** |
|---|---|---|
| **Cluster 1** | 111 (49%) | 17 (33%) |
| **Cluster 2** | 57 (25%) | 16 (35%) |
| **Cluster 3** | 42 (18%) | 3 (6%) |
| **Cluster 4** | 19 (8%) | 13 (27%) |

**Legend:** Significance assessed with Chi-squared test (p<0.001).

**eTable 12: Random Forest Classifier**

| | Predicted: | | | |
|---|---|---|---|---|
| Observed: | **Cluster 1** | **Cluster 2** | **Cluster 3** | **Cluster 4** |
| **Cluster 1** | 115 | 12 | 0 | 0 |
| **Cluster 2** | 8 | 51 | 10 | 5 |
| **Cluster 3** | 1 | 7 | 37 | 0 |
| **Cluster 4** | 5 | 10 | 0 | 17 |

**Legend:** Derivation Cohort confusion matrix of actual cluster assignments vs. those predicted by the random forest classifier using out-of-bag (OOB) predictions.

**eTable 13: Validation Cohort Characteristics (Utrecht, The Netherlands)**

| **Demographics (n=57 patients)** | |
|---|---|
| Age (median years, IQR) | 3.1 (IQR 1.3-13.5) |
| Sex (male) | 33 (57.9%) |
| Race<br>- Caucasian<br>- African/North African<br>- Middle Eastern<br>- Asian/SE Asian<br>- Eastern European/Russian<br>- Multiracial/Other | <br>42 (80.1%)<br>5 ( 9.6%)<br>2 ( 3.9%)<br>1 ( 1.9%)<br>1 ( 1.9%)<br>1 ( 1.9%) |
| **Medical History (n=57 patients)** | |
| Disease<br>- Leukemia[a]<br>- Inborn errors of immunity[b]<br>- Inborn errors of metabolism[c]<br>- Non-malignant hematologic[d]<br>- Lymphoma[e] | <br>24 (42.1%)<br>14 (24.6%)<br>13 (22.8%)<br>5 ( 8.8%)<br>1 ( 1.7%) |
| HCT Type<br>- Allogeneic<br>- Bone marrow<br>- Peripheral blood<br>- Umbilical cord blood (UCB) | <br>57 (100.0%)<br>- 13 (22.8%)<br>- 4 ( 7.0%)<br>- 40 (70.2%) |
| HLA match<br>- Matched related donor (BM/PB only)<br>- Matched unrelated donor (BM/PB only)<br>- Mismatched unrelated donor (BM/PB only)<br>- Matched UCB<br>- Mismatched UCB | <br>6 (10.5%)<br>10 (17.5%)<br>1 ( 1.7%)<br>15 (26.3%)<br>25 (43.9%) |
| Conditioning Regimen[f]<br>- Alkylating agent<br>- Busulfan<br>- Cyclophosphamide<br>- Melphalan<br>- Etoposide<br>- Treosulfan<br>- Antimetabolite<br>- Clofarabine<br>- Fludarabine<br>- Serotherapy (ATG or Alemtuzumab)<br>- Total body irradiation (TBI) | <br><br>49 (86.0%)<br>15 (26.3%)<br>3 ( 5.3%)<br>7 (12.3%)<br>1 ( 1.8%)<br><br>9 (15.8%)<br>41 (71.9%)<br>40 (70.2%)<br>6 (10.5%) |
| **Characteristics at time of BAL (n=57 patients)** | |
| Days from HCT to BAL (n, %) | 70 (IQR 21-104) |
| Comorbidities prior to BAL (n, %)<br>- acute GVHD preceding BAL<br>- any grade<br>- grade 3 or 4<br>- chronic GVHD preceding BAL<br>- any stage<br>- extensive | <br><br>22 (38.6%)<br>7 (12.3%)<br><br>11 (19.3%)<br>4 ( 7.0%) |

| | |
|---|---|
| - VOD preceding BAL<br>- any severity | 6 (10.5%) |
| BAL Clinical Microbiology Results (n, %)[g]<br>- Any positive<br>- Bacterial<br>- Viral<br>- Fungal/Protozoal<br>- More than 1 organism | <br>23 (40.4%)<br>4 ( 7.0%)<br>18 (31.6%)<br>12 (21.1%)<br>10 (17.5%) |
| **Outcomes at 365 days post BAL (n=57 patients)** | |
| Survival to last follow-up<br>Non-relapse mortality<br>Relapse | 31 (52.6%)<br>19 (35.1%)<br>7 (12.3%) |

**Legend:** [a] includes ALL (n=13), AML (n=10), JMML (n=1). [b] includes SCID (n=3), HLH (n=2), CGD (n=4), WAS (n=1), other (n=4). [c] includes Hurler syndrome (n=6), metachromatic leukodystrophy (n=3), other (n=4). [d] includes SAA (n=2), Fanconi anemia (n=1), thalassemia (n=1), other (n=1). [e] includes Hepatosplenic T-cell lymphoma (n=1). [f] Patients may have received multiple agents in the same or multiple categories. [g] Bacteria included Moraxella catarrhalis (n=1), Mycobacterium kansasii (n=1), Staphylococcus aureus (n=1), Stenotrophomonas maltophilia (n=1). Viruses included Adenovirus (n=5), CMV (n=1), Coronavirus (n=3), EBV (n=1), HHV-6 (n=3), HSV-2 (n=1), Metapneumovirus (n=1), Parainfluenzavirus 2 or 4 (n=1), RSV (n=1), Rhinovirus (n=11). Fungi/protozoa included Aspergillus (n=7 by positive galactomannan but no culture growth, n=2 by culture growth but negative galactomannan), Candida (n=1), Cladosporium (n=1), Pneumocystis (n=1).

**eTable 14: Validation Cohort Cluster Assignments**

| BAL Classification | N=57 | Average Ratio of Assigned Cluster vs Cluster 1 | Average Ratio of Assigned Cluster vs Cluster 2 | Average Ratio of Assigned Cluster vs Cluster 3 | Average Ratio of Assigned Cluster vs Cluster 4 |
|---|---|---|---|---|---|
| Cluster 1 | 21 | -- | 3.4 | 28.9 | 5.1 |
| Cluster 2 | 11 | 2.6 | -- | 7.2 | 2.9 |
| Cluster 3 | 24 | 4.4 | 1.3 | -- | 6.6 |
| Cluster 4 | 1 | 2.3 | 1.3 | 13.8 | -- |

**Legend:** Random forest classifier for BAL cluster, assigned to validation cohort. Final assignments determined as most likely outcome from applying random forest. Ratio of assignments to designated class vs each other class was computed for each of the n=57 patients in the validation cohort, and the mean ratios across each assigned cluster are listed.

**eFigure 1: HCT Day and Immune Counts at Time of BAL**

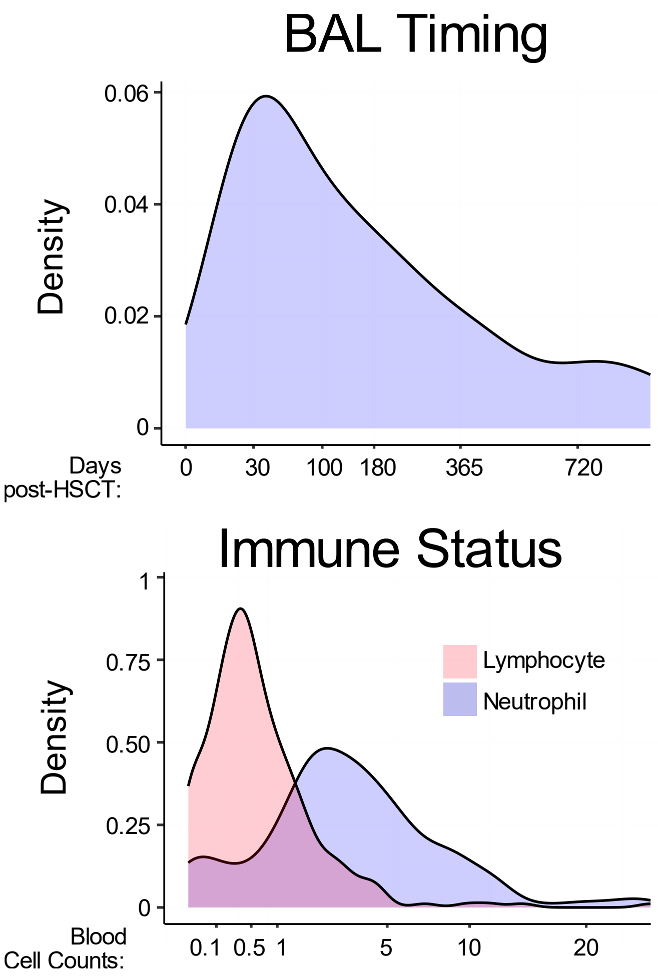


**Legend:** Kernel density plot of timing of BAL relative to HCT and blood neutrophil and lymphocyte counts prior to BAL**.**

**eFigure 2: MOFA Factor Correlation**

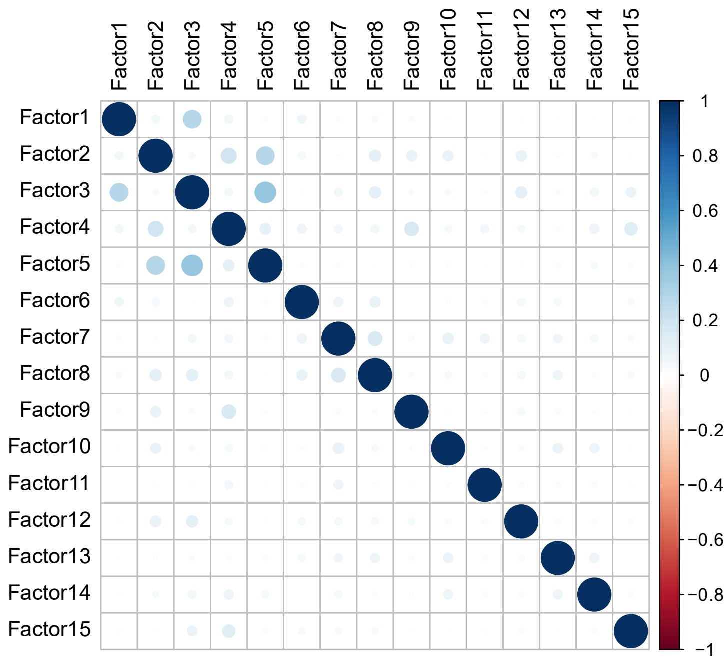


**Legend:** Fifteen latent factors were calculated for each patient, using all available microbiome and gene expression data. Latent factors showed minimal cross-correlation.

**eFigure 3: Percent of Variance Explained by MOFA Factors**

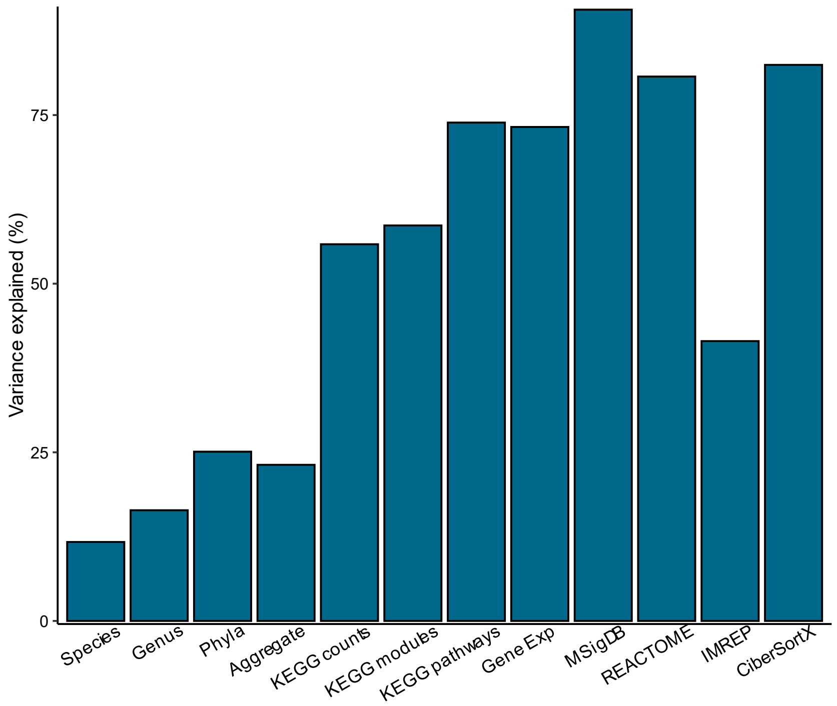


**Legend:** The overall variance explained by the sum of all fifteen latent factors. Greater total variance was explained for lung gene expression variables than for microbiome variables.

### eFigure 4: Selection of Optimal Number of Clusters

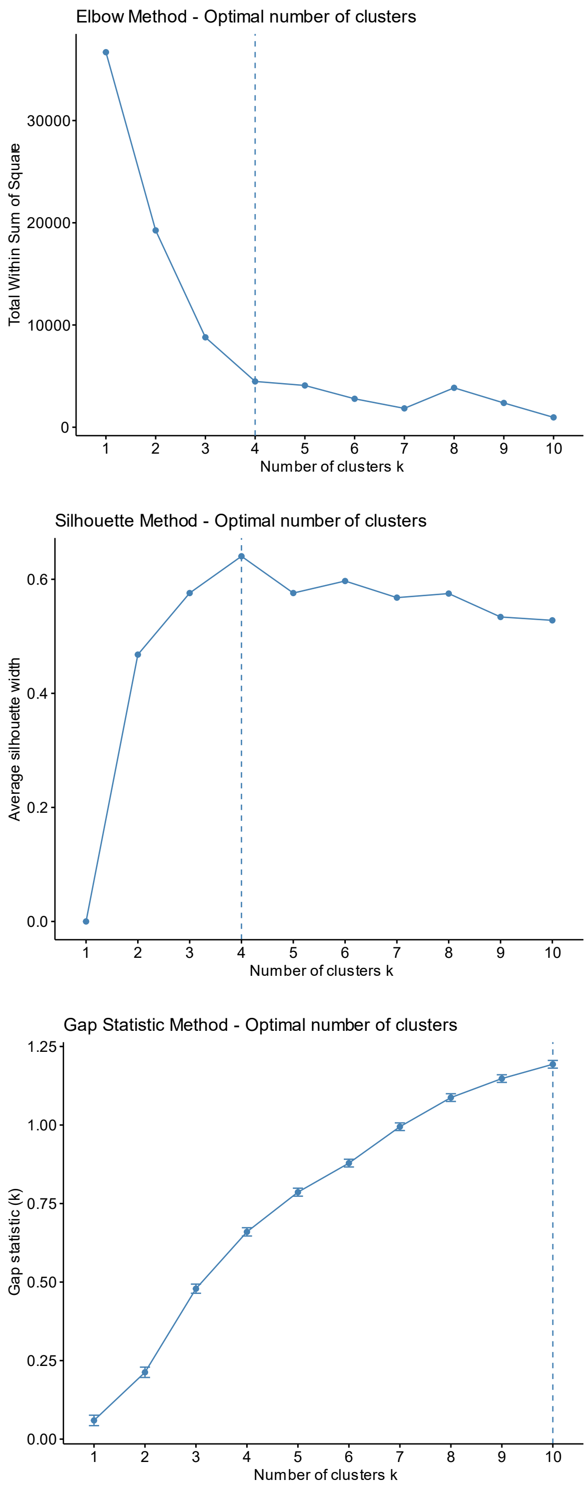


**Legend:** Elbow plot, silhouette plot, and gap statistic plot depicting ideal number of clusters based on umap of MOFA factors.  Based on these metrics, 4 clusters were chosen.

**eFigure 5: Representative BALs from Each Cluster**

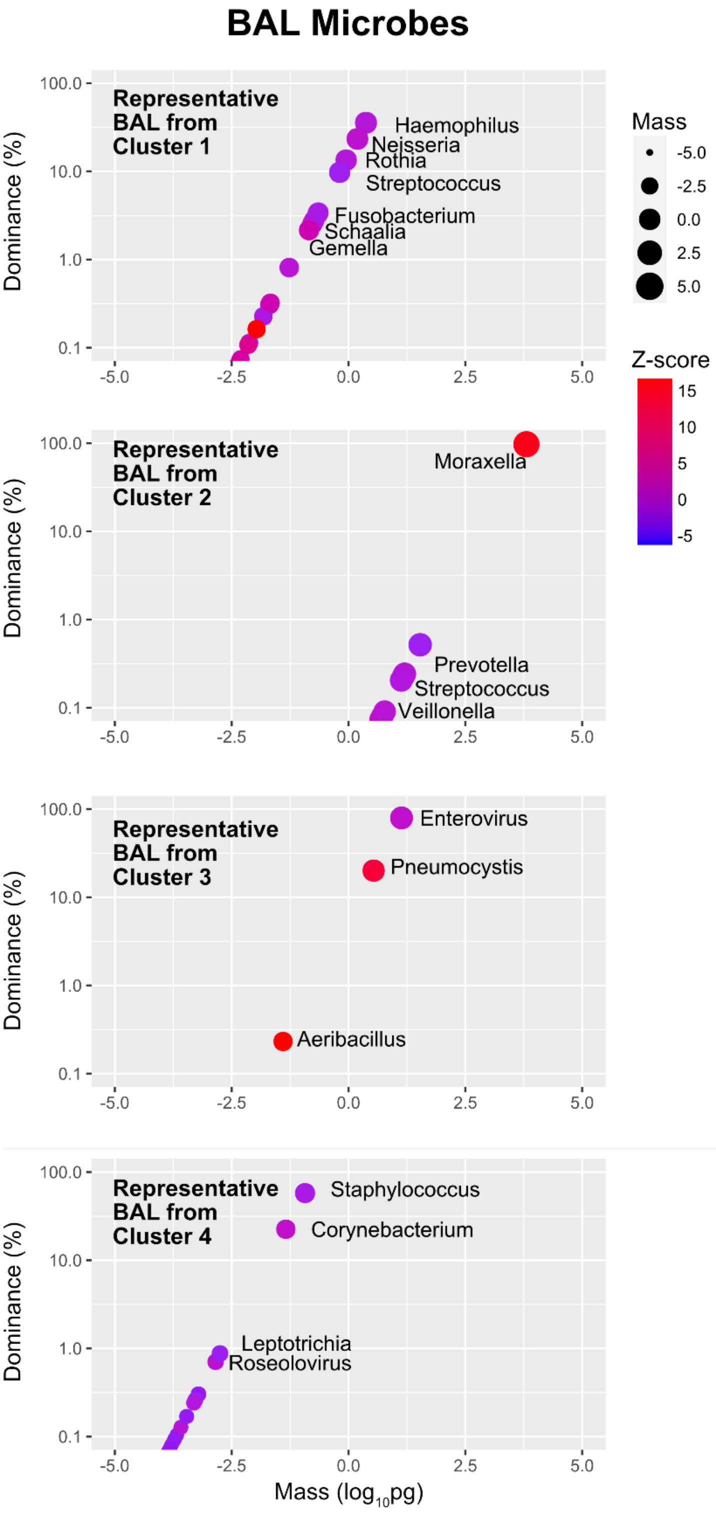


**Legend:** Representative BALs from each cluster are shown to illustrate example taxonomic compositions.

**eFigure 6: BAL Microbial Expression of KEGG for Carbohydrate, Energy, and Lipid metabolism pathways.**

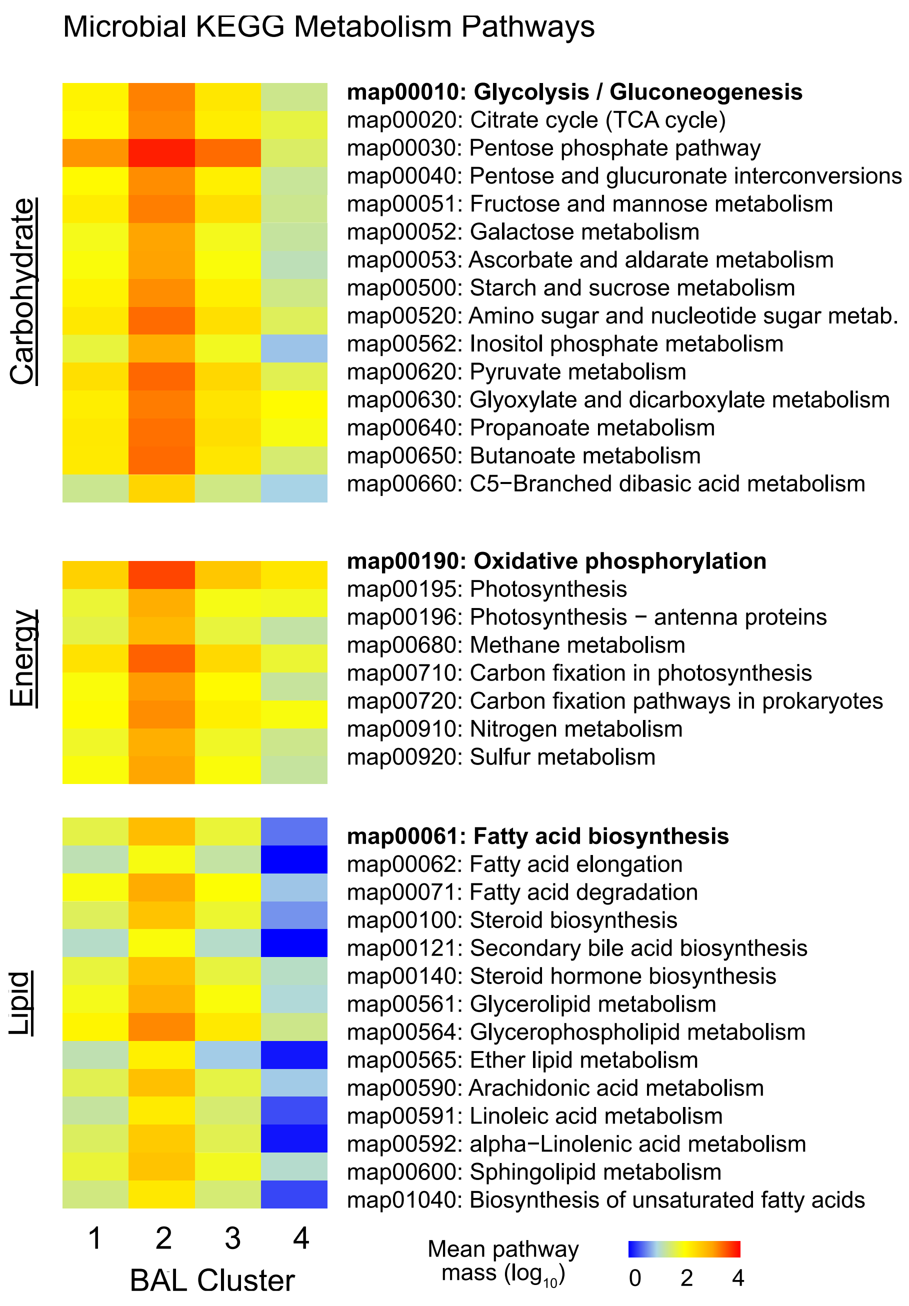


**Legend:** Mean ERCC-transformed normalized KEGG pathway expression for Carbohydrate, Energy, and Lipid metabolism pathways.

**eFigure 7: BAL Microbial Expression of KEGG Glycan Biosynthesis Pathways**

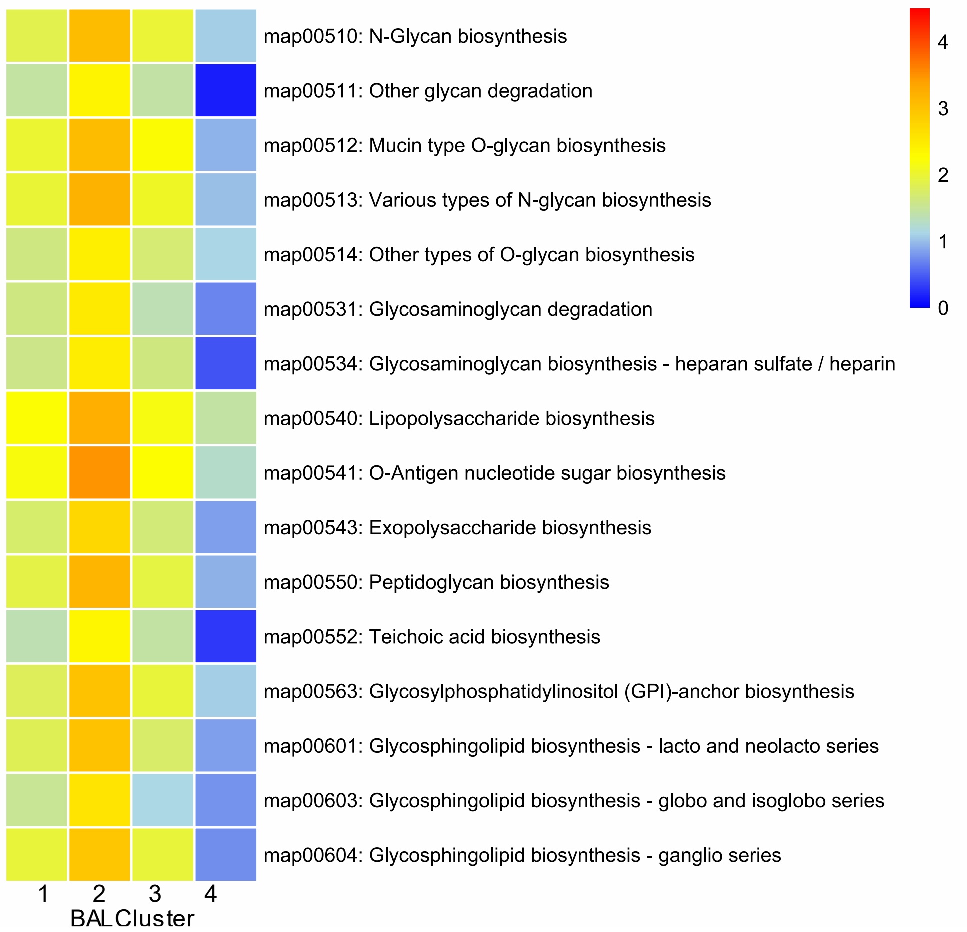


**Legend:** Mean ERCC-transformed normalized KEGG pathway expression for Glycan Biosynthesis Pathways.

**eFigure 8: In-Hospital Survival Stratified by Metagenomics Results**

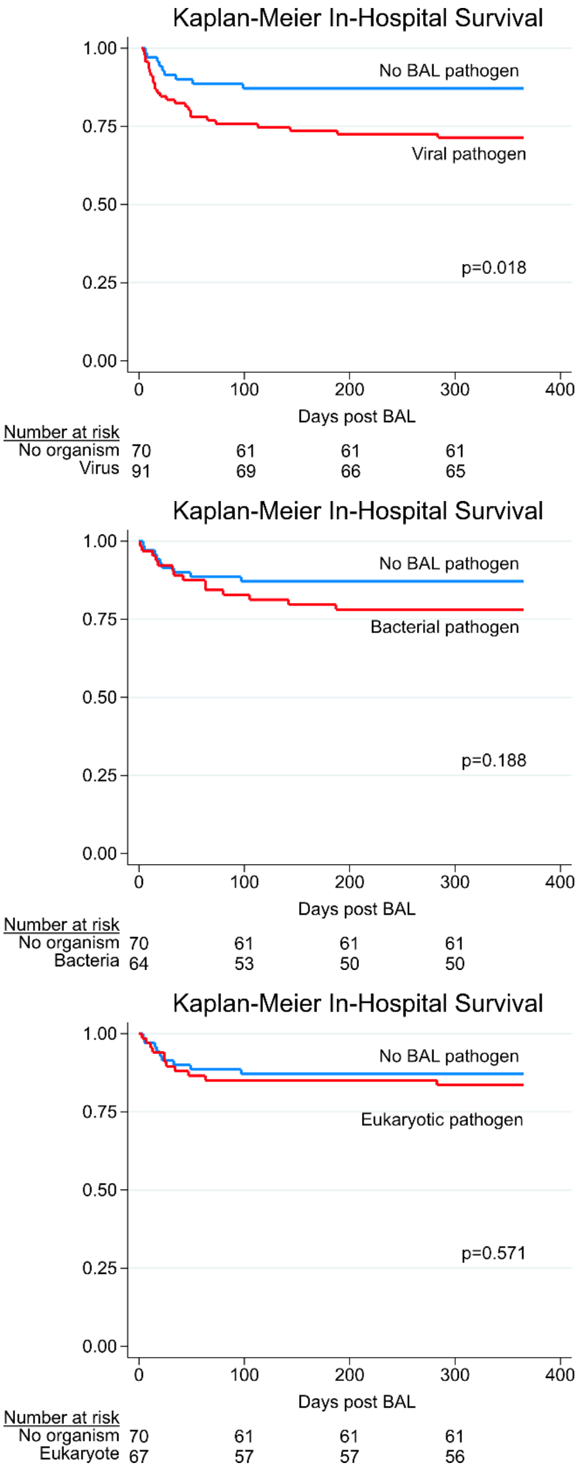


**Legend:** In-hospital survival for patients with a viral pathogen (top), bacterial pathogen (middle), or eukaryotic pathogen (bottom) detected on BAL, relative to no pathogen detected on BAL.

**eFigure 9: In-Hospital Survival Stratified by Antibacterial Exposure Score**

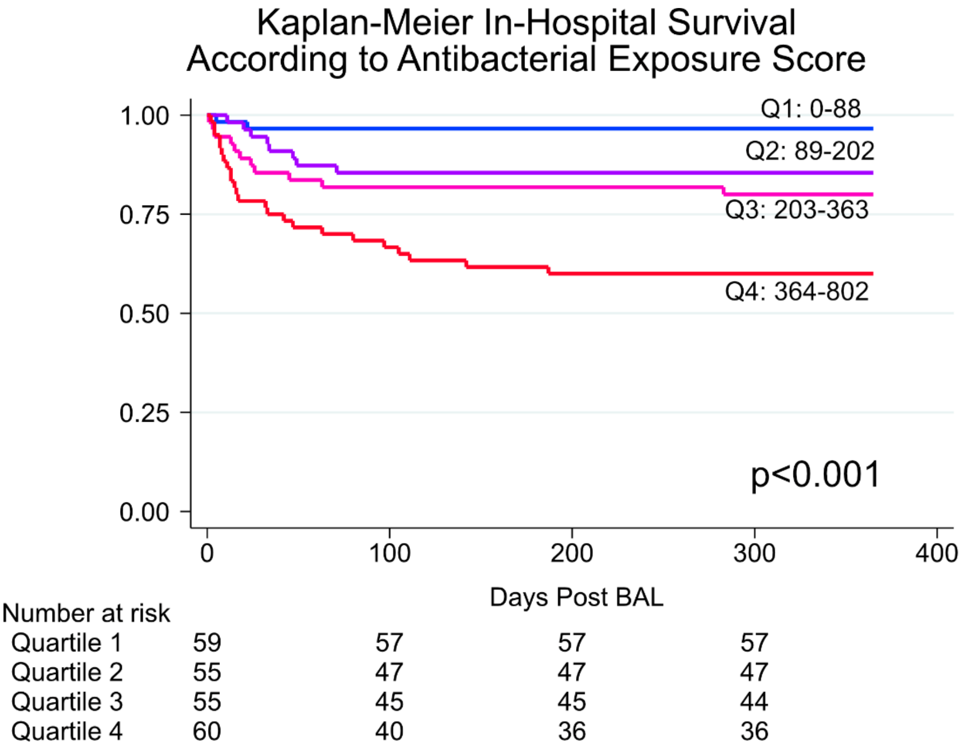


**Legend:** Antibacterial exposure score (AES) was divided into 4 quartiles of equal patient number and in-hospital survival was plotted for each quartile and compared with the log-rank test.

**eFigure 10: Causal Mediation of Effect of Antibiotic Exposure on Outcome**

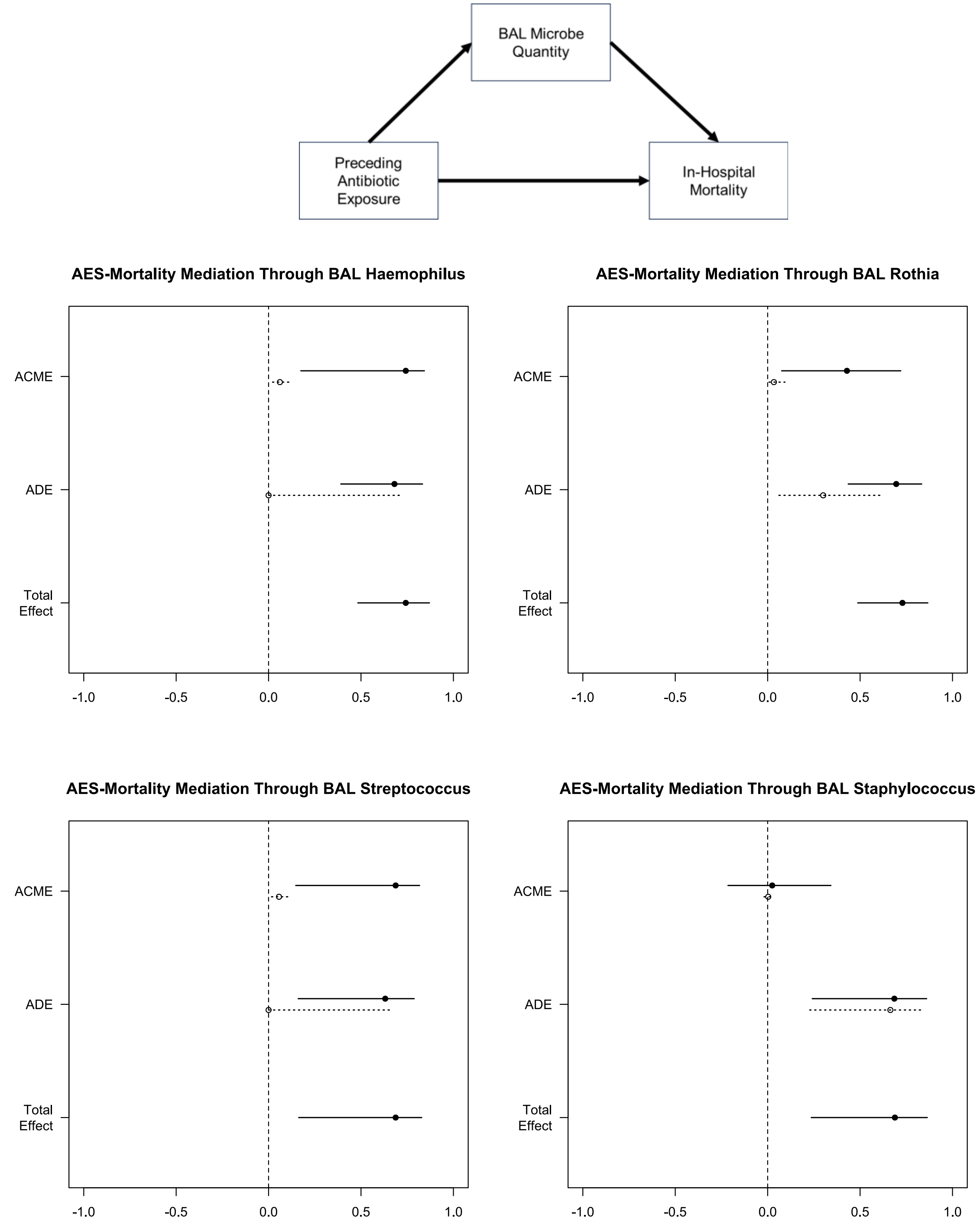


**Legend:** Causal Mediation plots indicating total effect of AES on in-hospital mortality, with average direct effects (ADE) and average causal mediation effects (ACME) plotted for treated (solid) and untreated (dotted) patients. For modeling purposes, treated refers to an AES of 800, and untreated refers to an AES of 0.

**eFigure 11: Imputed BAL Cell Fractions**

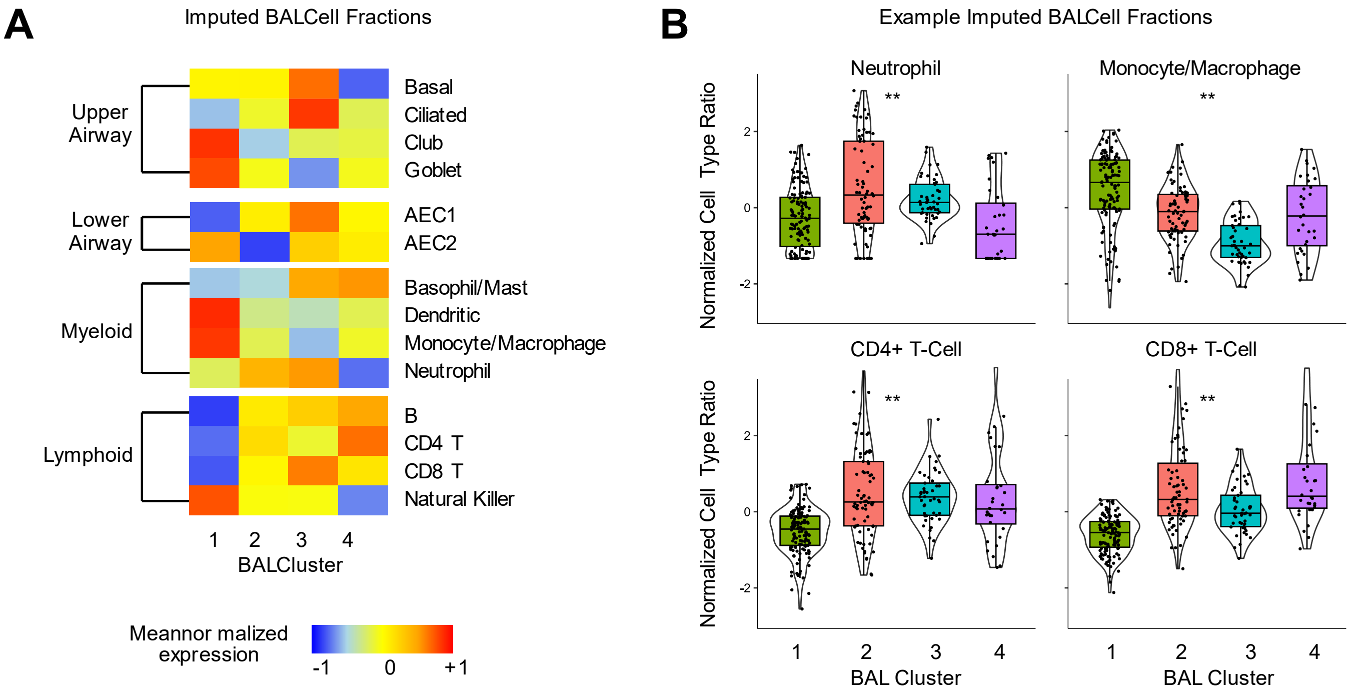


**Legend:** Cell type fractions were imputed using bulk gene expression and a reference single cell lung atlas. **(A)** Mean centered, scaled fractions are depicted for each cluster. **(B)** Raw values are shown for specific cell types.

**eFigure 12: Imputed Monocyte/Macrophage-Specific Gene Expression**

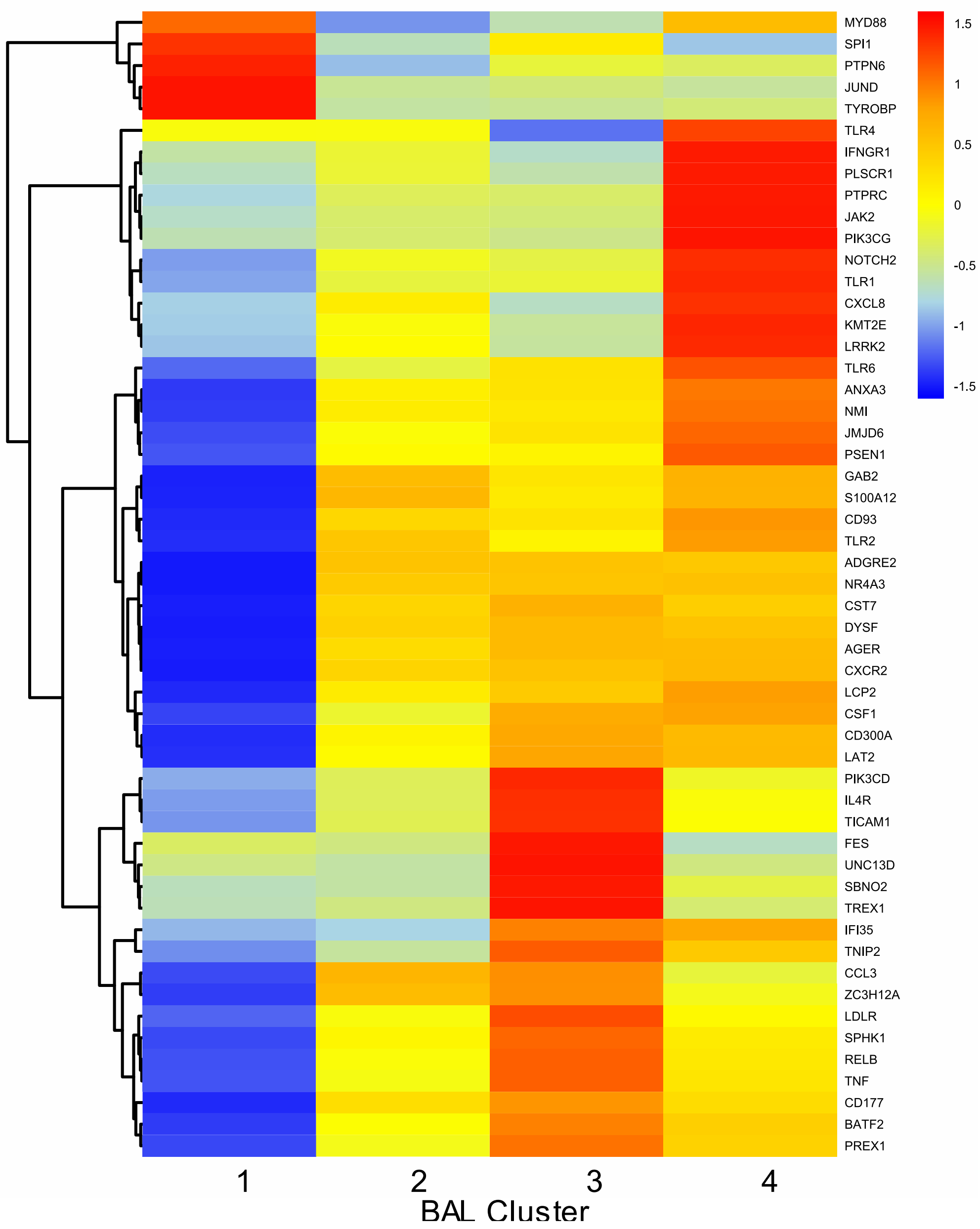


**Legend:** Monocyte/macrophage-specific expression of the "GOBP Myeloid Leukocyte Activation" gene set was imputed. Genes that were statistically significantly differentially expressed across clusters were selected for the heatmap, and average cell-type specific gene expression across the 4 clusters is displayed.

**eFigure 13: Imputed Lymphocyte-Specific Gene Expression**

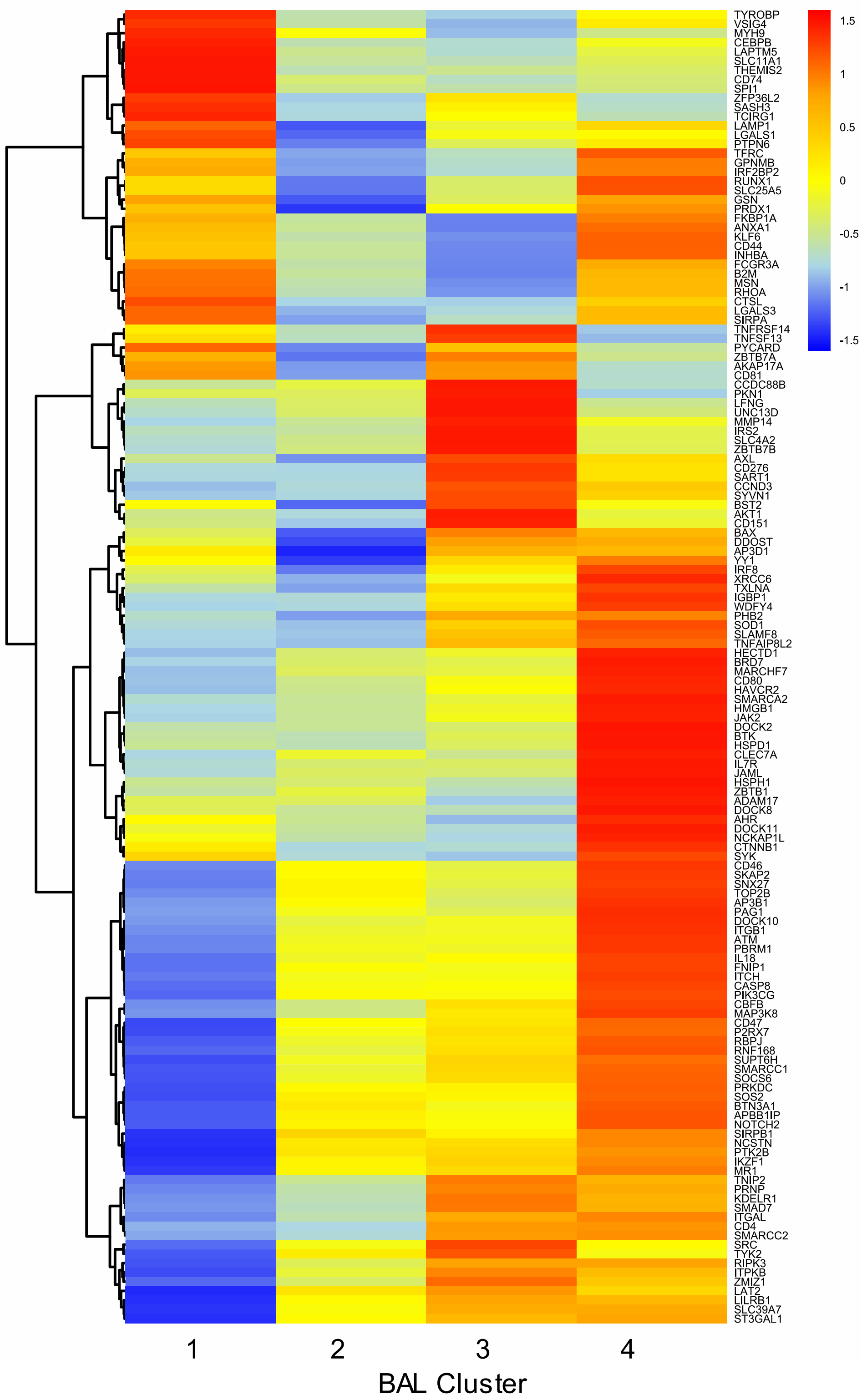


**Legend:** Lymphocyte-specific expression of "GOBP Lymphocyte Activation" gene set was imputed. Genes that were statistically significantly differentially expressed across clusters were selected for the heatmap, and average cell-type specific gene expression across the 4 clusters is displayed.

**eFigure 14: T-Cell Receptor Alpha Reads**

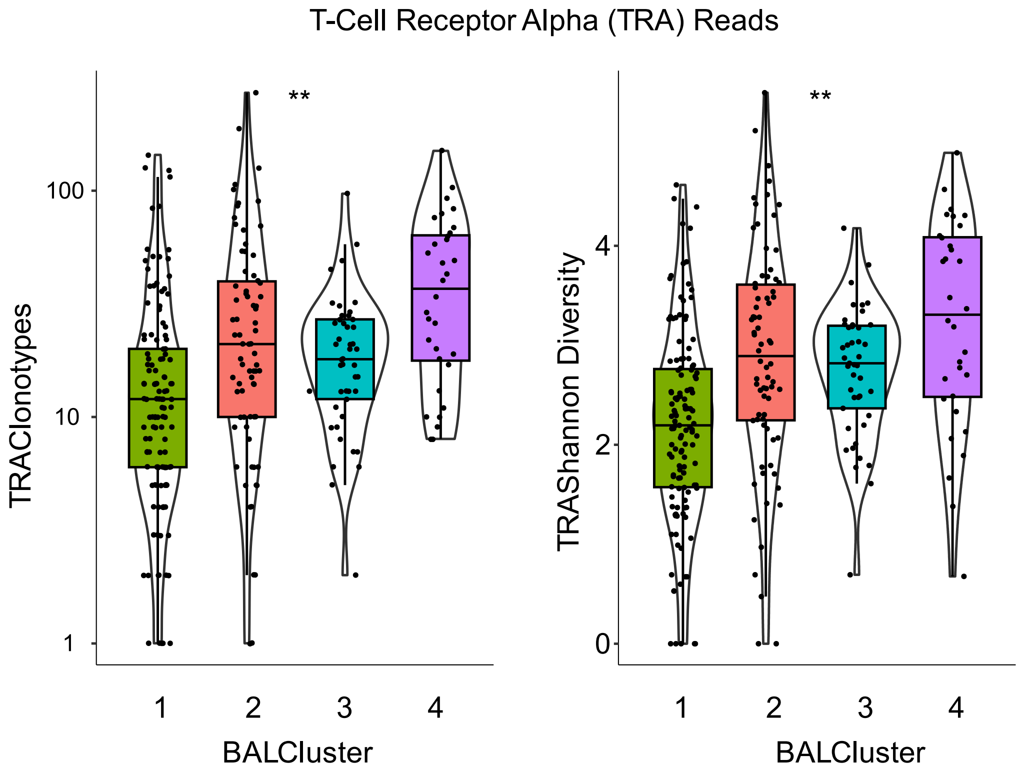


**Legend:** CDR3 alignments were computed and clonotypes and Shannon diversity of TCRα alignments are shown for each of the BAL clusters.

**eFigure 15: BAL Cluster Transitions**

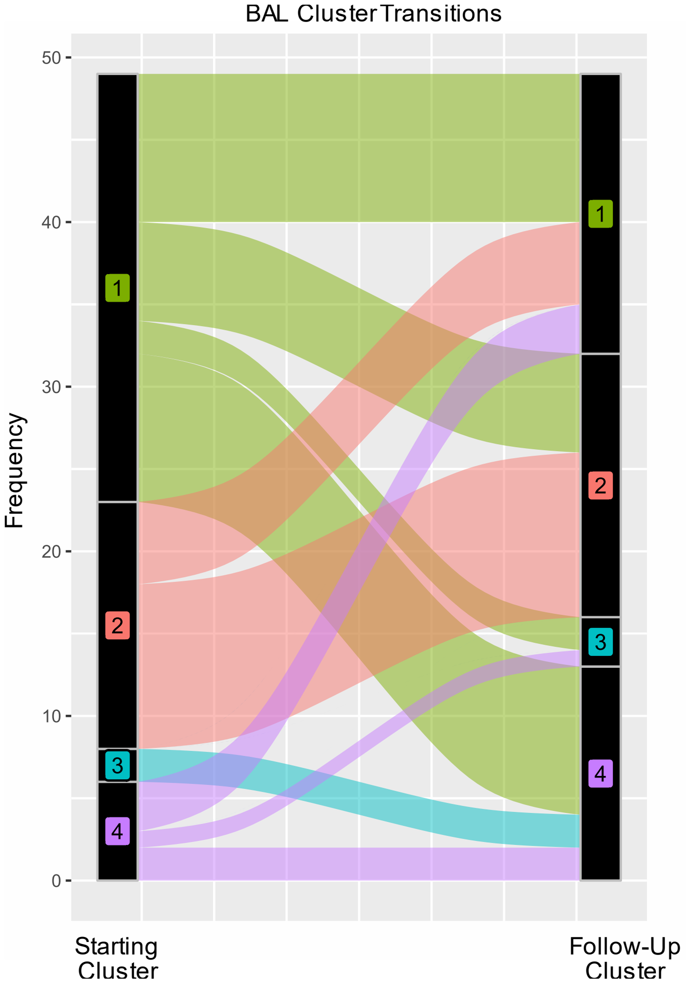


**Legend:** 34 patients had ≥2 BALs in this study (total 49 BALs were repeat samples). Cluster transitions are shown here, indicating a general transition away from the low-risk Cluster 1 on repeat samples.

**eFigure 16: Validation Cohort Survival Curves**

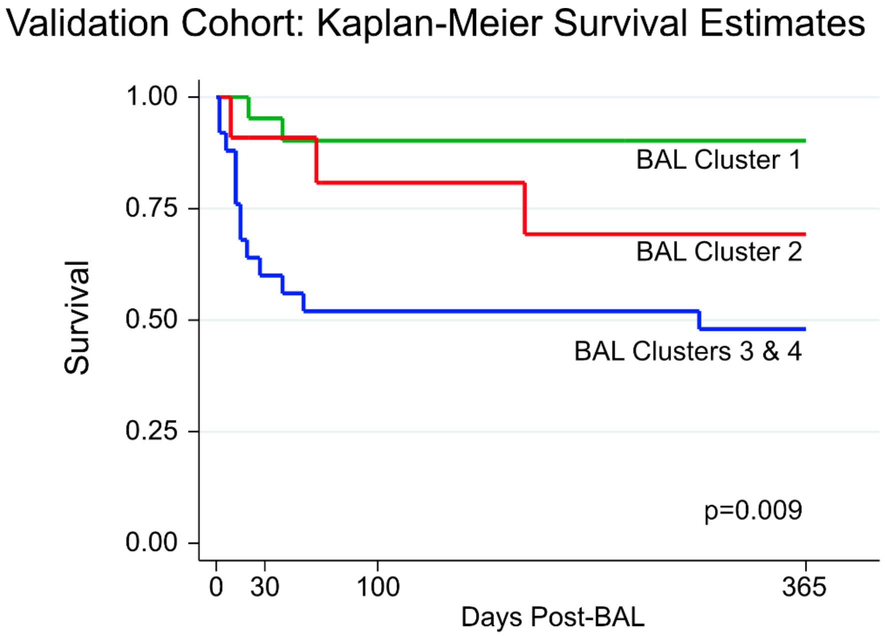


| Number at risk | 0 | 30 | 100 | 365 |
|---|---|---|---|---|
| Cluster 1 | 21 | 19 | 18 | 17 |
| Cluster 2 | 11 | 9 | 7 | 6 |
| Cluster 3 or 4 | 25 | 13 | 13 | 12 |

**Legend:** A random forest classifier using BAL metagenomic and transcriptomic data was grown using the derivation validation set. The classifier was applied to BAL data from a validation cohort, and 1-year non-relapse mortality was plotted according to cluster assignment and compared using the log-rank test.
